# Mild HIV-specific selective forces overlaying natural CD4+ T cell dynamics explain the clonality and decay dynamics of HIV reservoir cells

**DOI:** 10.1101/2024.02.13.24302704

**Authors:** Daniel B. Reeves, Danielle N. Rigau, Arianna Romero, Hao Zhang, Francesco R. Simonetti, Joseph Varriale, Rebecca Hoh, Li Zhang, Kellie N. Smith, Luis J. Montaner, Leah H. Rubin, Stephen J. Gange, Nadia R. Roan, Phyllis C. Tien, Joseph B. Margolick, Michael J. Peluso, Steven G. Deeks, Joshua T. Schiffer, Janet D. Siliciano, Robert F. Siliciano, Annukka A. R. Antar

**Affiliations:** Vaccine and Infectious Diseases Division, Fred Hutchinson Cancer Center, Seattle WA 98144 USA; Department of Global Health, University of Washington, Seattle WA 98144 USA; School of Medicine, Johns Hopkins University, Baltimore, MD 21205 USA; Department of Molecular Microbiology and Immunology, Bloomberg School of Public Health, Johns Hopkins University, Baltimore, MD 21205 USA; School of Medicine, University of California San Francisco, San Francisco, CA 94158 USA; Bloomberg Kimmel Institute for Cancer Immunotherapy, Johns Hopkins University, Baltimore, MD 21205 USA; The Wistar Institute, Philadelphia, PA 19104 USA; Gladstone Institutes, San Francisco, CA 94158 USA; Department of Medicine, University of Washington, Seattle WA 98144 USA; Howard Hughes Medical Institute, Baltimore, Maryland, USA

**Keywords:** HIV cure, HIV reservoir, latent reservoir, mathematical modeling, ecology, TCR, clonal, antigen-specific, CD4+ T cells, reservoir decay

## Abstract

The latent reservoir of HIV persists for decades in people living with HIV (PWH) on antiretroviral therapy (ART). To determine if persistence arises from the natural dynamics of memory CD4+ T cells harboring HIV, we compared the clonal dynamics of HIV proviruses to that of memory CD4+ T cell receptors (TCRβ) from the same PWH and from HIV-seronegative people. We show that clonal dominance of HIV proviruses and antigen-specific CD4+ T cells are similar but that the field’s understanding of the persistence of the less clonally dominant reservoir is significantly limited by undersampling. We demonstrate that increasing reservoir clonality over time and differential decay of intact and defective proviruses cannot be explained by mCD4+ T cell kinetics alone. Finally, we develop a stochastic model of TCRβ and proviruses that recapitulates experimental observations and suggests that HIV-specific negative selection mediates approximately 6% of intact and 2% of defective proviral clearance. Thus, HIV persistence is mostly, but not entirely, driven by natural mCD4+ T cell kinetics.

## Introduction

HIV persists in people living with HIV (PWH) on antiretroviral therapy (ART) as proviruses integrated primarily in the genomes of CD4+ T cells^1–3^. Most proviruses are genome defective, i.e., are not replication competent or intact^4,5^. HIV proviruses that are replication competent comprise the HIV latent reservoir^6,7^, which persists for decades^8–14^ and can seed rebound viremia if ART is stopped for any reason^15–18^. Thus, the reservoir is the major barrier to cure of HIV^19,20^.

There is strong evidence that reservoir persistence in blood and tissues during ART is due to cellular proliferation of infected CD4+ T cells. Heavy-water (deuterium) labeling experiments in PWH on ART have demonstrated that median CD4+ T cell half-lives are generally much shorter than the half-life of intact proviruses *in vivo*^5,23–25^, suggesting that proliferation of CD4+ T cells repopulates the reservoir. Providing direct evidence of this, replication competent proviruses with identical genomes and identical integration sites have been isolated from sister cells of clonal populations^26–33^. Model-based extrapolations of HIV proviral sequence data suggest that reservoirs are extremely clonal – after one year on ART, over 99% of CD4+ T cells harboring proviruses have divided at least once and thus are members of a clonal population^34^. These clones are not all the same size, however. Observational and modeling work demonstrate that the reservoir contains a small number of highly expanded or dominant clones and a massive number of smaller clones^34,35^.

The frequency of cells harboring replication competent^8,9^ proviruses among all CD4+ T cells declines over the first decade after ART initiation^10–14^. Cells harboring defective proviruses also decline, but at a slower rate^5,10,13,14,36^. Therefore, some cells disappear and what remains in the reservoir has been observed to become increasingly concentrated in highly expanded clones over time on ART^13,14,27,37,38^. Modeling of these observational data suggests that after a decade on ART, over half of an individual’s remaining proviruses are members of the top 100-1000 most-highly expanded infected clones^13^.

It remains unknown whether the natural drivers of CD4+ T cell proliferation and death in humans (e.g., antigen stimulation and homeostatic proliferation among others) entirely account for the extraordinary clonality of HIV proviruses and their decay dynamics. Antigen-driven clonal proliferation of HIV-infected cells could certainly account for many or most of the highly expanded proviral clones^39–41^. The role of homeostatic proliferation in the maintenance of the reservoir is less clear. Factors related to provirus sequence^42,43^ or integration site^26,27,37,44,45^ may confer selective advantages or disadvantages to certain proviral clones and therefore contribute to the reservoir’s extraordinary clonality. While provirus integration into oncogenes is now thought to play only a minor role in reservoir clonal expansion^46^, there is evidence that integration into transcriptionally inactive heterochromatin may confer a survival advantage^44,47–50^. The largest infected clones in PWH on ART with low-level viremia are more consistently present in short-term sampling compared to uninfected memory CD4s of the same size, perhaps reflecting different proliferative drivers or HIV-specific selective advantages^35^.

To test whether the natural physiology of memory CD4+ T cells (e.g., HIV as inert “passenger” hypothesis) is sufficient to explain reservoir clonality and dynamics or whether selective factors related to HIV infection itself are required to explain these observations, we generated longitudinal clonality data from memory CD4+ T cells in HIV seronegative individuals and from HIV proviruses and CD4s simultaneously from PWH on ART. Specifically, we investigated whether the passenger hypothesis could explain three key observations: (1) the high degree of HIV proviral clonality in PWH on ART, (2) the increase in proviral clonality over time on ART, and (3) the differential decay of cells harboring intact and defective proviruses over time on ART. We compared the longitudinal (∼10 year) clonality of HIV proviruses (intact and defective) from resting CD4+ T cells with the longitudinal clonality of their putative carrier cells, resting memory CD4+ (rmCD4) T cells from the same blood samples from PWH on ART. We also compared proviral clonality of the replication competent reservoir at single time points from PWH on long-term ART with the clonality of rmCD4s from the same blood samples. To determine whether suppressed HIV infection itself is associated with significantly different mCD4 clonality, we compared the longitudinal cohort of PWH on ART to HIV-seronegative people matched on age, race, and sex. We identified antigen specific (CMV and HIV) mCD4 cells in a subset of people and compared their clonality and decay rates with that of HIV-infected cells and all mCD4s. To make these comparisons, we modeled clonotype abundance distributions and calculated clonality and diversity metrics. Finally, we developed a mechanistic mathematical model of CD4+ clonotypes in which some harbor HIV proviruses (intact and defective). We matched this model to our comprehensive experimental data and arrived at the conclusion that mild HIV-dependent forces such as negative selection on mostly intact HIV proviruses are required to account for the increasingly oligoclonal composition of the reservoir and its decay over time on ART.

## Results

### Participants and sampling

We sampled people enrolled in four cohorts: two longitudinal cohorts (4 PWH on ART and 4 HIV-seronegative people) and two cross-sectional cohorts (7 PWH on ART and 33 HIV-seronegative people) (**Table 1, Figure 1A**, detailed information in **Table S1 & Figure S1**).

**Figure 1.**
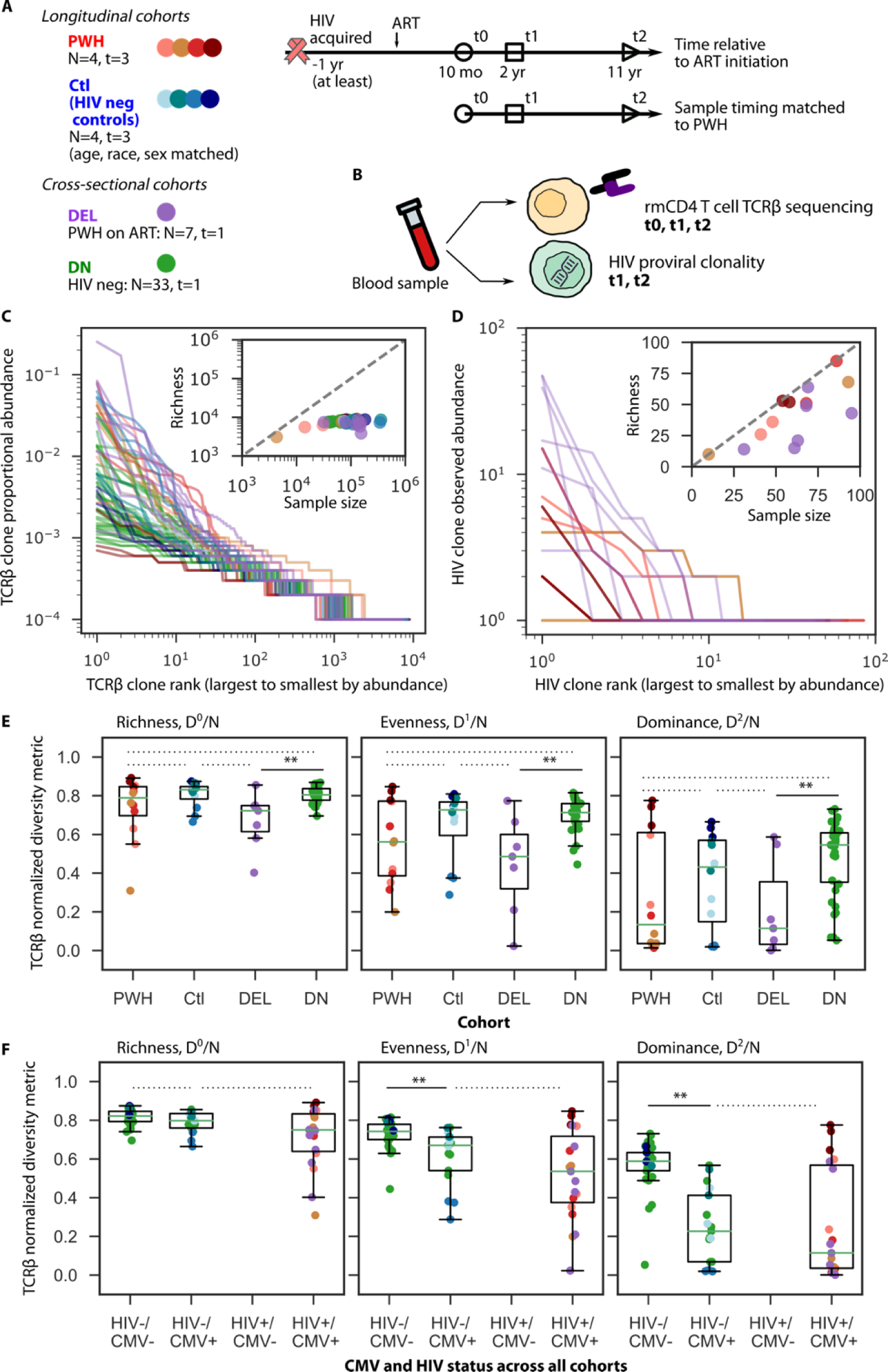
HIV-infected cell and resting memory CD4+ T cell clonality in PWH and HIV-seronegative individuals. A) Four PWH (**Table 1**) were sampled longitudinally at 3 time points on average 10 months, 2 years, and 11 years after ART initiation. Four age-, race-, and sex-matched HIV-seronegative individuals were sampled at the same intervals. Seven PWH on ART and 33 HIV-seronegative individuals were sampled at one timepoint. B) TCRβ repertoires were used to assess clonality of mCD4 cells in all participants. HIV provirus sequences and env sequences from replication competent outgrowth viruses were used to assess clonality of HIV in PWH. C) mCD4 clonality is shown as rank abundance curves, in which clonotypes are arranged from most to least abundant along the x axis (colors from **A** denote cohorts). Inset: Observed richness, or number of unique clonotypes, deviates from the line y=x, saturating at higher sample sizes. D) HIV clone rank abundance at absolute abundance observed and colored by cohort. Inset: Observed richness roughly follows the line y=x, increasing with increasing sample sizes. E) Ecological diversity metrics (Hill numbers D^X^/N calculated from data resampled to the same size and normalized by dividing by the common sample size N=10^4^) calculated for mCD4+ TCRβ repertoires by cohort. F) The same ecological metrics stratified for all cohorts by HIV and CMV serostatus. Horizontal lines in **E** and **F** indicate comparisons tested with 1 sided Mann-Whitney test, ** above solid lines indicates p<0.005 and dashed lines indicate p>0.05.

**Table 1.** Summary of longitudinal and cross-sectional cohorts. Detailed information for all participants is available in Supplementary Table 1.

| Name | Type | HIV status | Time points | N | Sex | Age range at t0 | t0 | t1, t2 | TCRβ cell type | HIV cell type (sequence) |
| --- | --- | --- | --- | --- | --- | --- | --- | --- | --- | --- |
| **PWH** | Longitudinal | Pos | 3 | 4 | 3M<br>1F | 35-40 | TCR | TCR/<br>HIV | rmCD4 | rCD4 (nfl seq, total proviruses) |
| **Control (Ctl)** | Longitudinal | Neg | 3 | 4 | 3M<br>1F | 36-39 | TCR | TCR | rmCD4 | N/A |
| **DELPSC**<br>**(DEL)** | Cross-sectional | Pos | 1 | 7 | 7M<br>0F | 41-60 | TCR/<br>HIV | N/A | rmCD4 | rCD4 (*env* seq<br>from QVOA+<br>wells) |
| **De**<br>**Neuter**<br>**(DN)** | Cross-sectional | Neg | 1 | 33 | 11M<br>22F | 21-50 | TCR | N/A | mCD4 | N/A |

Abbreviations: PWH, people living with HIV; TCRβ , T cell receptor $\beta$ chain; Pos, positive; Neg, negative; N, number of participants; M, male; F, female; N/A not applicable; rmCD4, resting memory CD4+ T cells; mCD4, memory CD4+ T cells; rCD4, resting CD4+ T cells; nfl, near-full length HIV provirus; seq, sequences; QVOA+, quantitative viral outgrowth assay positive, meaning virions were produced; env, HIV envelope

#### 1) Longitudinal PWH on ART (PWH): HIV provirus vs rmCD4 clonality

To investigate the clonality of HIV-infected cells, we examined near-full length (nfl) HIV-1 provirus sequences (intact and defective) from peripheral resting CD4+ T cells (mean 57 sequences per time point, IQR 46-73) from four PWH^11^ who remained on ART for over ten years with no documented lapses in therapy after ART initiation and with undetectable viral loads (aside from 3 blips) checked generally every six months or more frequently (**Figure S1, Methods**). Each participant had 3 samples collected at an average of 10 months (t0), 2 years (t1) and 11 years (t2) after ART initiation (**Figure 1A-B**)^13^. Proviruses (nfl) were sequenced at t1 and t2, and rmCD4+ TCRβ repertoires were sequenced at t0, t1, and t2 (sorting schema in **Figure S2**).

#### 2) Longitudinal HIV-seronegative people (Control, Ctl): rmCD4 clonality

As controls for our longitudinal PWH cohort, we obtained longitudinal samples from four confirmed HIV-seronegative people from the MACS/WIHS combined cohort study who were originally enrolled based on having similar risk behaviors as enrolled PWH^51^. These four were matched one-to-one to the four longitudinally sampled PWH on ART described above on age, race, sex, time between samples, and CMV seropositive status (with one exception, **Table S1**). rmCD4s were sorted and TCRβ repertoires were obtained.

#### 3) Cross-sectional PWH on ART (DELSPC, DEL): Reservoir clonality vs rmCD4 clonality

To investigate the clonality of cells comprising the replication competent reservoir, clonality was approximated using *env* sequences from outgrowth viruses from quantitative viral outgrowth assays performed at limiting dilution (QVOAs) from one sample an average of 12 years (IQR 9-15 yr) after ART initiation from a separate cohort of 7 PWH on ART^52^. rmCD4s were sorted and TCRβ repertoires were obtained from the same samples.

#### 4) Cross-sectional HIV-seronegative people (De Neuter, DN): mCD4 clonality

As controls for our cross-sectional PWH cohort, we obtained single-timepoint memory CD4+ TCRβ repertoires from a cohort of 33 HIV-seronegative individuals^53^.

### Clonality of HIV-infected vs mCD4+ T cells

CD4+ T cell clonality (m or rmCD4) was approximated using TCRβ DNA sequencing repertoires^54^ (average 1.3 x 10^5^ TCRs per sample, **Table S1**, sorting schema in **Figure S2, Methods**).

HIV-infected cell clonality in the longitudinal cohort was approximated by identifying cells harboring proviruses with identical nucleotide sequences and, if deleted, with identical deletion junctions, as has been done previously^13,55,56^. The clonality of HIV reservoir cells in the cross-sectional PWH cohort was approximated by identifying outgrowth viruses harboring identical *env* sequences, as has been done previously^57^. HIV sequence sample sizes are limited due to the rarity of HIV-infected cells in PWH on ART and the labor-intensive nature of near-full length provirus sequencing and viral outgrowth assays^4^. Because the vast majority of rmCD4s are not infected with HIV, these experiments are in effect comparing the clonality of HIV-uninfected CD4s to that of HIV-infected cells, as has been done previously^35^.

### rmCD4 and HIV clonality both follow a characteristic log-log linear rank-abundance distribution

We first used rank-abundance distributions to enable visual and semi-quantitative comparisons of clonality (**Figure 1C-D**). In these plots, each clonotype is “ranked” or ordered (x-axis) by its observed abundance (y-axis) from most highly expanded (x=1) to least. Clonotypes that have the same observed abundance are randomly ranked. A log-log scale emphasizes the large numbers of unique clonotypes on the x-axis (richness) as well as the orders of magnitude differences in abundances on the y-axis. rmCD4 proportional rank abundance curves generally followed a characteristic log-log linear shape across all participants living with and without HIV and across all time points, with varying slopes (**Figure 1C**). Similar to our previous observations^13,34^, rank abundances for HIV sequences, including for replication competent sequences, also followed a similar overall shape, though curves are substantially less smooth because of lower sample sizes (**Figure 1D**).

### Richness of circulating memory CD4+ cell TCRβ clonotypes saturates with increasing sample size

Richness, a term for the number of unique clonotypes found in a sample, is one simple measure of clonality/diversity. Although mCD4 TCRβ sample sizes were heterogeneous across individuals (**Table S1**), we observed that sequence richness did not increase indefinitely with sample sizes and instead approximates 10^4^ across all PWH and HIV-seronegative person-time points (**Figure 1C inset**). This implies a general richness, or a count of unique clonotypes, of mCD4s on the order of 10^4^ in sample sizes of less than 1 million total circulating mCD4s, in line with prior observations from HIV-seronegative people^58^. In contrast, the richness of HIV sequences generally increased with increasing sample sizes (**Figure 1D inset**), implying that sampling depths of up to 100 HIV-infected cells per time point do not come close to detecting most HIV-infected clonotypes in the peripheral circulation.

### Quantitative comparisons of clonality require resampled diversity metrics

Ecological diversity metrics have been used to summarize the clonality of HIV-infected cells in the HIV literature^34^. Common metrics include richness, the Shannon index^59^, a measure of clonal unevenness, and the Simpson index^60^, a measure of concentration into dominant clonal groups. Here, we use normalized Hill numbers, which are transformations of these 3 metrics into a mathematically unified and more interpretable family of indices (**Supplementary Methods**)^61^. Hence, we use the total number of unique clonotypes (Richness, D^0^), the exponential of the Shannon index which approximates the relative evenness of all clonotypes (Evenness, D^1^), and the inverse of the Simpson index which approximates the relative dominance of the largest clonotypes (Dominance, D^2^).

These Hill numbers are also sensitive to sample sizes^62^. By assuming an underlying distribution that was more or less clonal, we showed normalized Hill numbers decrease when the same distribution is sampled with higher sample sizes. The sample size dependence also grew as the underlying true distribution became more clonal (**Figure S3**). Hence, to make all comparisons we computationally resample data to the same size (**Figure S3A**), and only then compute normalized Hill numbers on equally sampled data. In conclusion, we report Hill numbers after resampling equally, and then divide by the sample size such that each metric D^X^/N is comparable across participant time points and ranges from 0 (highly clonal) to 1 (highly diverse).

### Minimal differences in mCD4 cell TCRβ clonality across cohorts

There were no significant differences in rmCD4 cell TCRβ clonality between the PWH on ART and HIV-seronegative longitudinal cohorts that were matched for age, race, sex, and CMV status (**Figure 1E**). The cross-sectional PWH (DEL) cohort had generally more clonal rmCD4 cells than the cross-sectional HIV-seronegative (DN) cohort for all three diversity metrics, despite the fact that DN includes actively proliferating (non-resting) CD4+ T cells. This discrepancy could be due to differences in ages and health status between the cross-sectional cohorts. For instance, DEL participants are significantly older than DN participants (mean age 51 vs 37, p=0.001, 1 sided Mann-Whitney U test) (**Table S1**).

### Cytomegalovirus (CMV) infection but not suppressed HIV infection is associated with the clonality of mCD4+ T cell compartment

While it is well known that CMV infection in humans is associated with increased T cell clonality^63–66^, it is not clear whether HIV infection is similarly associated with perturbed clonality in the memory CD4+ T cell compartment. When data was grouped by HIV and CMV serostatus, we observed that, regardless of the presence of suppressed HIV infection, cytomegalovirus (CMV) serostatus was highly associated with decreased diversity metrics for mCD4s. No significant differences were observed for richness, but mCD4 repertoires from HIV-/CMV+ individuals were less even (p=0.0007) and displayed more clone dominance (p=2.4e-7) than those from HIV-/CMV-individuals (**Figure 1F**). Importantly, among people with positive CMV serology, there were no significant differences in mCD4 cell diversity metrics between PWH on ART and HIV-seronegative individuals. These results suggest that CMV infection in humans is associated with significantly increased mCD4 clonality while suppressed HIV infection in CMV+ individuals is not.

### HIV-infected cells are more clonal than general memory CD4+ T cells even after controlling for lack of new HIV diversity during suppressive ART

Two steps were required to fairly compare the clonality of HIV-infected rCD4s, harboring either intact or defective proviruses, and rmCD4 cells from the same longitudinal peripheral blood samples. First, unlike the memory CD4+ TCRβ pool which continually gains diversity as naïve T cells encounter antigen and enter the memory pool, ART reliably blocks viral replication^34,67–69^, meaning few if any novel clonotypes enter the HIV-infected cell pool in PWH with suppressed viral loads on ART. HIV-infected CD4s have been observed to be more clonal than total memory CD4+ T cells from the same sample without controlling for this factor^40,71^. Therefore, we compared the clonality of HIV proviruses to “persistent TCRβ” clonotypes, which are TCRβs found at t1 or t2 that were also confirmed present at t0, a time point close to the initiation of ART (schematic in **Figure 2A, Figure 2B**).

**Figure 2.**
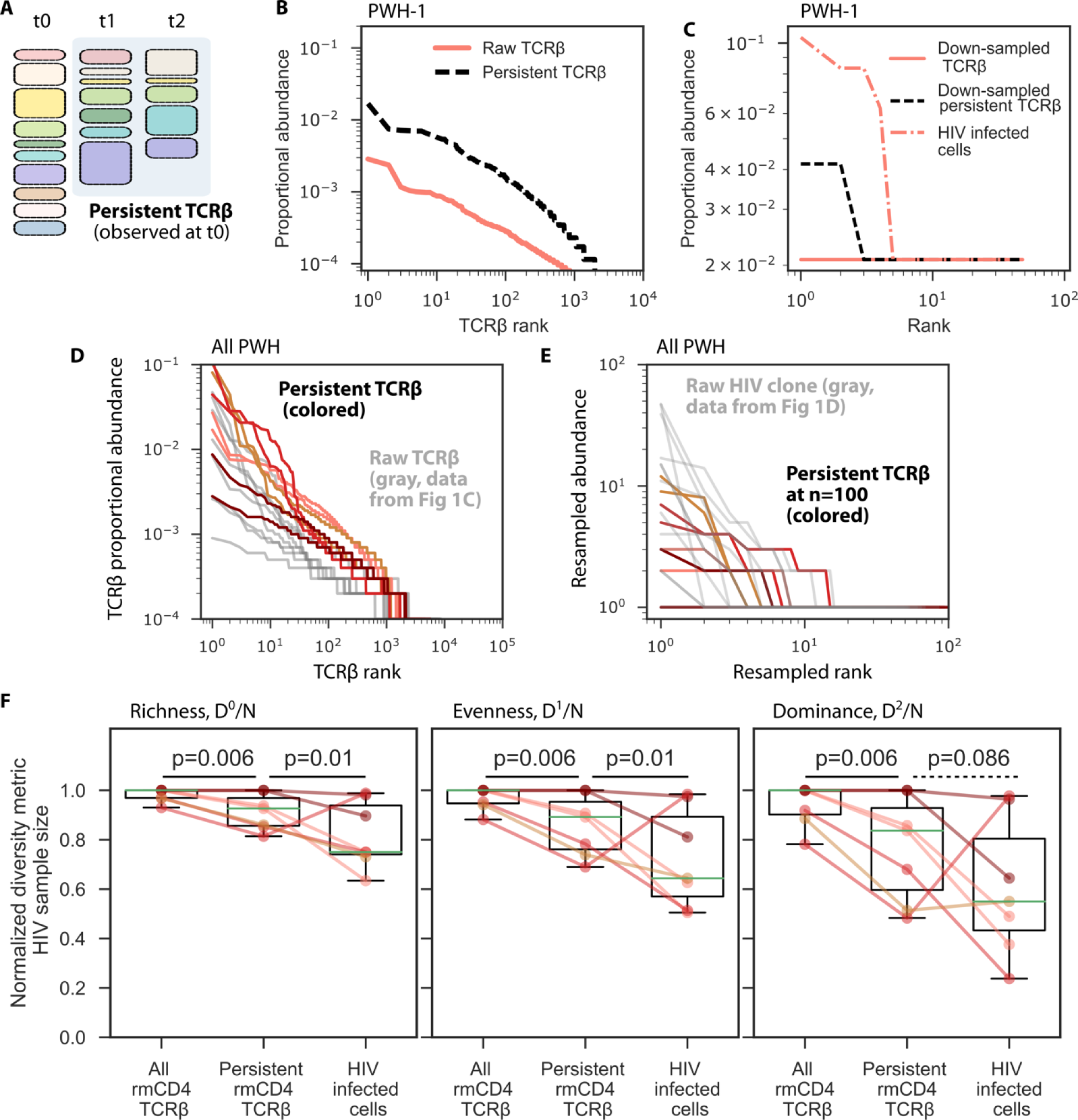
HIV proviruses are more clonal than persistent resting memory CD4+ T cells from the same peripheral samples. A) To fairly compare the clonality of HIV proviruses in PWH on ART to rmCD4 cells, which naturally gain new clonotypes over time as naïve T cells encounter antigen and enter the memory compartment, we define “persistent TCR” as TCRβs present at t1 and t2 that were also present in a sample close to the initiation of ART, t0. B) Proportional abundance of persistent TCRβ vs all raw rmCD4 clones for PWH-1(t1). C) Proportional abundances of downsampled TCRβ vs persistent TCRβ vs HIV proviruses for PWH-1(t1). Downsample size was set to the sample size of HIV proviruses from that participant time point (n=n_HIV_). D) All PWH persistent TCRβ (colored) vs raw TCRβ (gray) proportional rank abundances. E) All PWH persistent TCRβ downsampled to HIV sample size vs raw HIV-infected cell rank abundances. Rank abundances are statistically different, see **Figure S4. F**) Ecological diversity metrics for downsampled total rmCD4 cells, downsampled persistent TCRβ, and HIV proviruses from the seven (of eight) t1 and t2 PWH timepoints with n_HIV_>10. All downsample sizes were set to the sample size of HIV proviruses from that participant time point and metrics were normalized to this value. P-values indicate paired differences using a one-sided Mann-Whitney test.

Second, we computationally downsample persistent TCRβ clonotype distributions to the same sample size as HIV sequence sample sizes at the matching participant time point^46^. Rank abundance curves immediately suggest that HIV-infected cells are more clonal than persistent rmCD4s (**Figure 2C**).

Further, our ecological diversity metrics (Hill numbers, normalized by sample size) indicate that HIV proviruses are generally still more clonal than downsampled persistent TCRβ (p<0.05 for normalized richness and normalized exponential Shannon (*D*^0^/N and *D*^1^/N) metrics, **Figure 2D, Figure S4**). The normalized inverse Simpson metric (*D*^2^/N) was not significantly different, suggesting that despite differences in overall clonality, measures of clone dominance in rmCD4 and HIV proviruses are similar. Together, these analyses imply that even within the same individuals and even corrected for novel clonotypes (diversity) entering the rmCD4+ T cell compartment after ART initiation, HIV-infected CD4s in PWH on ART are generally more clonal and less diverse than the underlying population of persisting rmCD4+ T cells, though both contain dominant, highly expanded clones. The dominant, highly expanded clones may be responsive to common antigens across cohorts, such as CMV. To investigate the clonality of dominant clones further, we use a novel mathematical approach.

### Higher resolution rank abundance modeling reveals two potentially mechanistically different categories of resting memory CD4+ TCRβ clones

The incongruity between HIV-infected rCD4s and general rmCD4+ TCRβ clonality is interesting, given that mCD4+ T cells are the putative carrier cells for HIV proviruses, and that HIV clonality is generally understood to be driven by CD4+ T cell proliferation. Therefore, we refined our comparison using mathematical modeling of clonality distributions.

We previously modeled HIV sequence distributions using a power law model^34^ where abundance (*a*) follows rank (*r*) with *a*(*r*)∼*r*^−*α*^. This distribution suggests clonal abundances arise from stochastic dynamics where clonotypes have heterogeneous proliferation and death rates^70^. Meanwhile, the clonal structure of CD4+ T cells has also been modeled using a power-law distribution^71,72^. Therefore, we first tested our prior power law model on all mCD4+ TCRβ data (N=64 time points from 48 individuals, **Table S1**). Upon close inspection, we observed that rank abundances were not always precisely log-log linear. Instead, in some samples, there appeared to be two slopes, with a steeper slope for higher abundance clonotypes (examples in **Figure 3A**). To quantitatively test this hypothesis, we fit a double power law, or two-slope, model: *a*(*r*)∼*r*^−*α*_1_^ + *ϕr*^−*α*_2_^. Across all participants, the double power law model had lower root-mean-square (RMS) error, p=1e-5 (**Figure 3B**) and was a better fit to many distributions for both PWH on ART and HIV-seronegative individuals.

**Figure 3.**
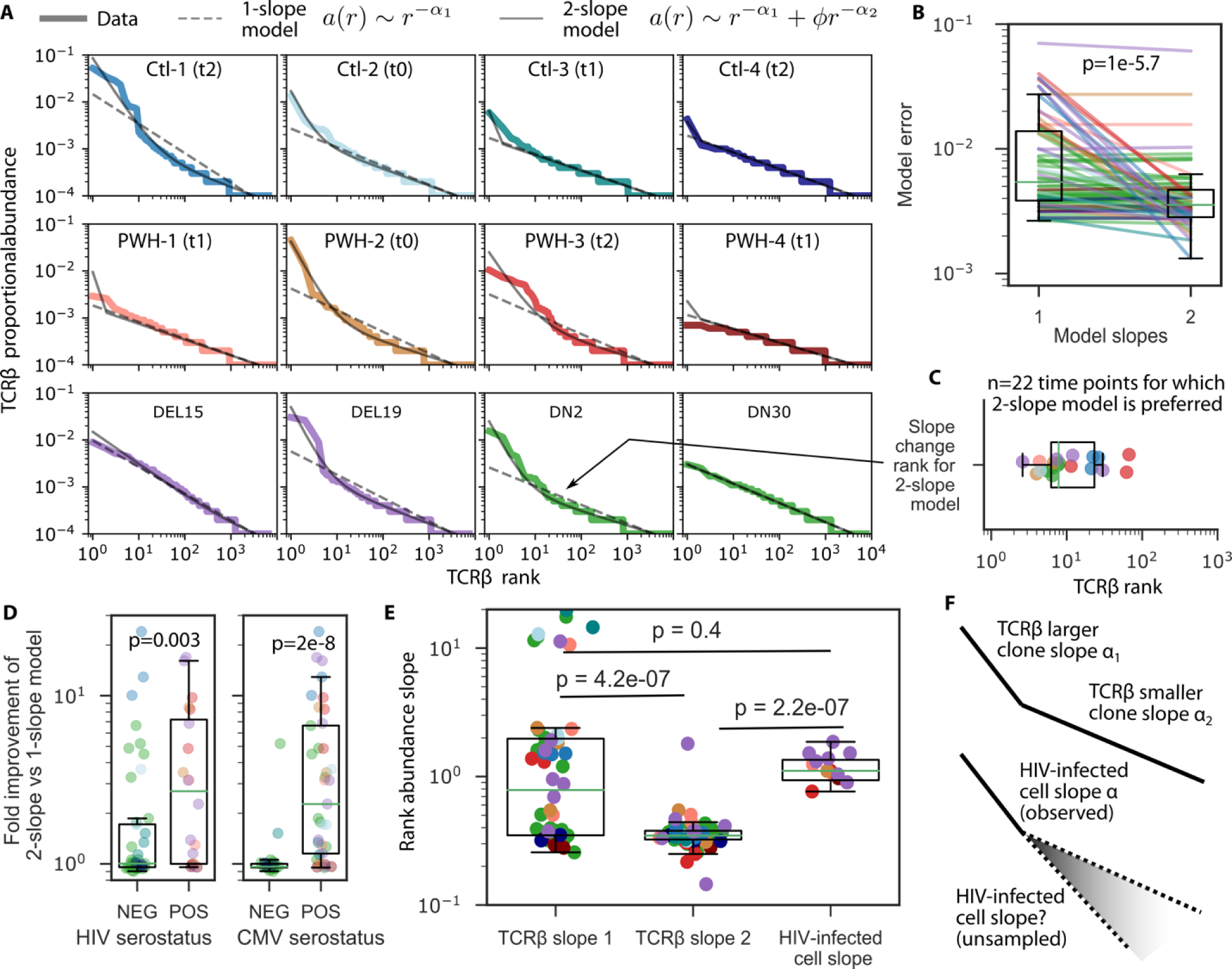
HIV infected cell clonality resembles that of the most highly expanded resting memory CD4+ T cell clones. A) Examples of fitting one slope and two slope power law models to rank abundances from PWH and HIV-negative individuals. B) Comparison of one and two slope model root-mean-square errors. C) The slope change rank, or the rank distinguishing the first and second slopes of the rank abundance curve. D) Fold improvement of model fit for HIV and CMV seropositive and seronegative individuals shows two-slope models are more likely (2-20 fold) to match data in seropositive individuals. E) Comparison of large and small TCRβ clone slopes to HIV-infected cell slopes. F) Cartoon illustration of the distribution shapes: HIV proviral clones and large TCRβ clones have similar slopes, but since sample size of HIV sequences is lower, we are not able to predict potentially smaller clone slopes in HIV clones. In all panels, p-values indicate 1 sided Mann-Whitney U test.

In addition, the two slopes in this model identify different regions of the rank abundance distribution. We calculated the “change point” or the rank distinguishing at which the slope changes (**Methods**). Among individuals for whom the two-slope model was preferred (2-fold improvement in fit error vs 1 slope model), the first slope pertained generally to the top 2-80 clonotypes, with median change point r=9 (**Figure 3C**). In other words, on average, the abundance of the top nine most highly expanded clones in a rmCD4 distribution were best modeled with a steeper power law slope than the rest of the clones. We hypothesized these larger clones might be driven by different dynamical mechanisms, such as recent antigenic response or inflationary memory, than the tens of thousands of smaller TCRβ clonotypes.

### CMV seropositivity associates with a two slope rmCD4+ T cell rank-abundance model

Some support for the hypothesis that clones in the larger of the two power law slope rank abundances are driven by antigenic expansions is generated by stratifying the individuals by HIV and CMV seropositivity. rmCD4+ TCRβ rank abundance distributions from HIV and CMV seropositive individuals were substantially more likely to require a two slope model than seronegative participants (p=0.003 and p=2e-8 for HIV and CMV, respectively, **Figure 3D**). The ratio of the error of the models showed the two slope model had 2-20x better fit for seropositive individuals whereas errors between one slope and two slope models were roughly equivalent for CMV seronegative, HIV seronegative participants. This finding implies that a steeper section of the rmCD4 clonotype rank abundance curve is sometimes found in HIV seropositive individuals and even more commonly found in CMV seropositive individuals, suggesting that the most highly expanded rmCD4 clonotypes could be responding to these viral antigens^73^.

We explored clinical variables including participant sex, age, months on ART, current CD4+ T cell count, nadir CD4+ T cell count, and estimated months of HIV infection before initiation of ART for associations with power law slope model parameters (**Figure S5**). After Bonferroni correction, significant associations were observed between male sex, positive CMV status, and low nadir CD4 count for the larger clone slope (*α*_1_), and between positive CMV status and low nadir CD4 count for the change point of the large and small clone slopes.

### rmCD4+ T cell large clone slope resembles HIV-infected CD4+ T cell clone slope

The first, or larger, clone slope from rmCD4s was significantly steeper than the smaller clone slope (p=1.3e-8, **Figure 3E**). More intriguingly, it was not significantly different from the HIV sequence rank abundance slope (p=0.2, **Figure 3E**), suggesting that the clonality of the most highly-expanded, ‘observed’ HIV proviruses may be driven by the same biological factors that drive the clonality of the most highly expanded rmCD4 clonotypes, e.g. antigen-specificity, effector ratio, lack of exhaustion^21,23,24,74^. The magnitude of the first slope indicates there are still large relative differences in clonal abundances; for example, the larger TCRβ clone slope of *α*_1_∼0.8 implies the 10^th^ largest clone is less than 1/6 the size of the largest clone. In contrast, the smaller TCRβ clone slope of *α*_2_∼0.4 (**Figure 3**) means the smaller clones are more similar in size; the 100^th^ clone may not be much smaller than the 10^th^, and the 1000^th^ is likely not much more than 1/10 the size of the 10^th^. Put another way, for clones modeled by the first slope, every 10-fold reduction in rank is associated with a 6-fold reduction in close size; for clones modeled by the second slope, every 10-fold reduction in rank is associated with a 2.5-fold reduction in clone size.

Although we can compare the clonality distributions of the largest rmCD4 clones and the largest HIV-infected clones, we cannot comment on similarities of smaller, less-expanded, HIV provirus clones to project whether they are best modeled by two slope power law models like rmCD4+ TCRβ clones, simply because the sampling depth of HIV-infected cells is not sufficient due to technical limitations of the assay (**Figure 3F**). Our observations imply that the mechanisms underlying the clonality of the largest observed HIV clones (e.g., antigen-driven stimulation) could be preferentially seen to maintain reservoirs by the HIV cure field, despite the possibility that other mechanisms (e.g. homeostatic proliferation) may maintain a larger in number, but less clonally dominant, population of reservoir cells.

Finally, we wanted to compare the clonality of the replication competent reservoir to all HIV-infected cells – which contain mostly defective proviruses. Indeed, we found DEL cohort participants had statistically higher HIV reservoir slopes compared to all proviruses from longitudinal PWH participants (**Figure S6**). This could be due to differences in cohort makeup, adherence patterns, time on ART, that DEL sequence clones are determined by HIV *env* sequencing vs near-full length sequencing, or because the replication competent reservoir truly is more clonal than all HIV-infected cells, but we could not distinguish these several factors.

### CMV-responding and HIV-responding rmCD4+ T cells have similar clonality to HIV infected cells

To address whether the most highly expanded rmCD4 clonotypes were specifically responding to two relevant and potentially common antigens in these participants, we identified CMV- and HIV-responding TCRβ sequences using the viral functional expansion of specific T cells (ViraFEST) assay from PWH-1 and −4, who were available to donate samples specifically for this assay at an additional time point t3 (16 and 19 years after ART initiation, **Figure 4A, Methods, Table S1**)^75^. The ViraFEST assay identifies TCRβ sequences of CD4+ T cells that proliferate *ex vivo* in response to antigen stimulation. We chose ViraFEST over other methods that identify cells expressing activation markers in response to antigen^76^ since we were interested in a clone’s ability to proliferate. We identified 335 CMV-specific and 69 HIV-specific TCRβ sequences from PWH-1 and 323 CMV-specific and 25 HIV-specific TCRβ sequences from PWH-4, which distributed throughout rank abundance distributions at all time points (**Figure 4B**). Across all participant time points, two CMV-specific clonotypes were found in the top 10 most highly expanded rmCD4 clonotypes and 24 CMV-specific clonotypes and 3 HIV-specific clonotypes were in the top 100 rmCD4 clonotypes.

**Figure 4.**
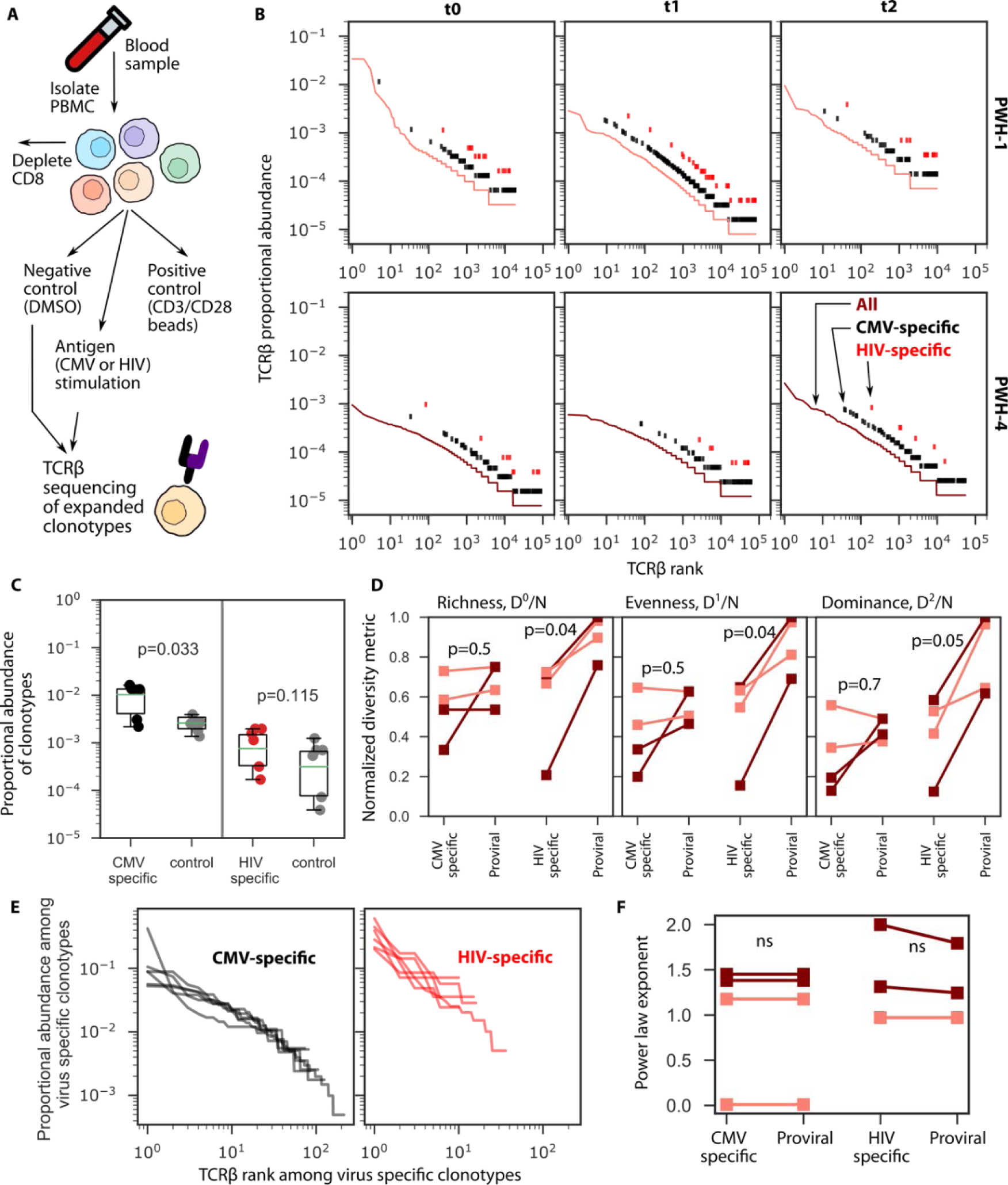
Clonality of CMV- and HIV-specific rmCD4+ T cells. A) Schematic of the ViraFEST assay, which identifies TCRβ sequences from CD4+ T cells that proliferate in response to specific antigens. B) rmCD4+ TCRβ rank abundance distributions for PWH-1 and PWH-4 (curves) with markers indicating the rank of clonotypes that proliferated in response to CMV (black) or HIV (red) antigen by ViraFEST at t3. C) Summed proportional abundances of antigen-specific clonotypes at each participant time point (red and black dots) vs proportional abundance of the same number of randomly selected rmCD4+ T cell clonotypes from the same person-time point. D) Normalized ecological diversity metrics comparing clonality of CMV-specific, HIV-specific, and HIV proviruses compared pairwise where each value in a pair is calculated by downsampling data to the lower sample size at each participant time point and normalizing metrics to that sample size. E) Rank abundance curves of only the CMV- and HIV-specific clonotypes, one curve per person-timepoint. F) Power law exponents of a one slope power law model of rank abundance curves of CMV-specific, HIV-specific, and HIV proviruses at each person-time point. In **D** and **F**, colors indicate PWH-1 (darker) and PWH-4 (lighter), p-values indicate paired t-test; ns, not significant p>0.05.

Across participant time points, CMV-specific rmCD4 clonotypes were significantly more abundant than the same number of randomly chosen rmCD4 clonotypes (p=0.03, **Figure 4C**. HIV-specific clonotypes were slightly more abundant than the same number of randomly chosen clonotypes but not significantly so. PWH-1 had rmCD4 distributions that required a two slope model, and the slope change ranks for this participant’s time points t0, t1, and t2 were φ=18, 3, and 10. One CMV-specific clonotype was observed in the first slope (at t0, **Figure 4B**) and proliferated *ex vivo* in response to antigen at t3, indicating that this CMV-specific rmCD4 clone has remained at high frequency in the peripheral circulation and retained its ability to proliferate in response to antigen over at least a 15 year timespan, in agreement with other work reporting on memory-inflated CMV-specific mCD4 clones over shorter time intervals in PWH who were not on ART^77^. The antigen specificity of the remaining clones in the first slope at each timepoint of PWH-1, remains unknown.

Using normalized diversity metrics, we compared the clonality of antigen specific rmCD4 cells to that of HIV-infected cells. Metrics were calculated by downsampling and normalizing to the lower sample size of the two types (antigen specific vs HIV-infected cells), and comparisons were made pairwise for each person-timepoint. CMV-specific rmCD4s were no more or less clonal than HIV proviruses (**Figure 4D**). HIV-specific rmCD4s were somewhat more clonal (p∼0.05) than HIV proviruses, but differences could be driven by the very low numbers of HIV-specific clones observed in 3 of 4 participant time points (N=198, 28, 14, and 29). Therefore, we examined power law exponents from rank-abundance distributions (**Figure 4E**) and observed they were not significantly different between antigen-specific cells and HIV proviruses (p>0.18, **Figure 4F**).

### The clonality of persistent and antigen specific rmCD4+ clones remains stable over time in people with and without HIV, whereas the clonality of HIV-infected CD4s increases over time within individuals

The characteristic shape of rmCD4 clonotype rank abundance curves for each longitudinal PWH on ART and HIV-negative participant did not have significant within-person differences across three samples spanning 10 years, although there are differences between people that are not attributable to HIV serostatus (**Figure 5A**). We previously showed that HIV-infected cells become concentrated in highly expanded clones over ∼10 years on ART in these individuals^13^ (**Figure S7**). By contrast, here we observe no similar increase in clonality in persistent rmTCRβ (**Figure 5B**) or all rmTCRβ (**Figure S8**) over ∼10 years on ART in the same individuals over the same time span. Αdditionally, although sample size was small, we also did not observe trends supporting changes in longitudinal clonality for CMV- and HIV-specific rmCD4+ T cells (**Figure S9**). These results agree with other demonstrations of the stability of clonally dominant CD4+ T cells over time spans of ∼ 2 years^78^.

**Figure 5.**
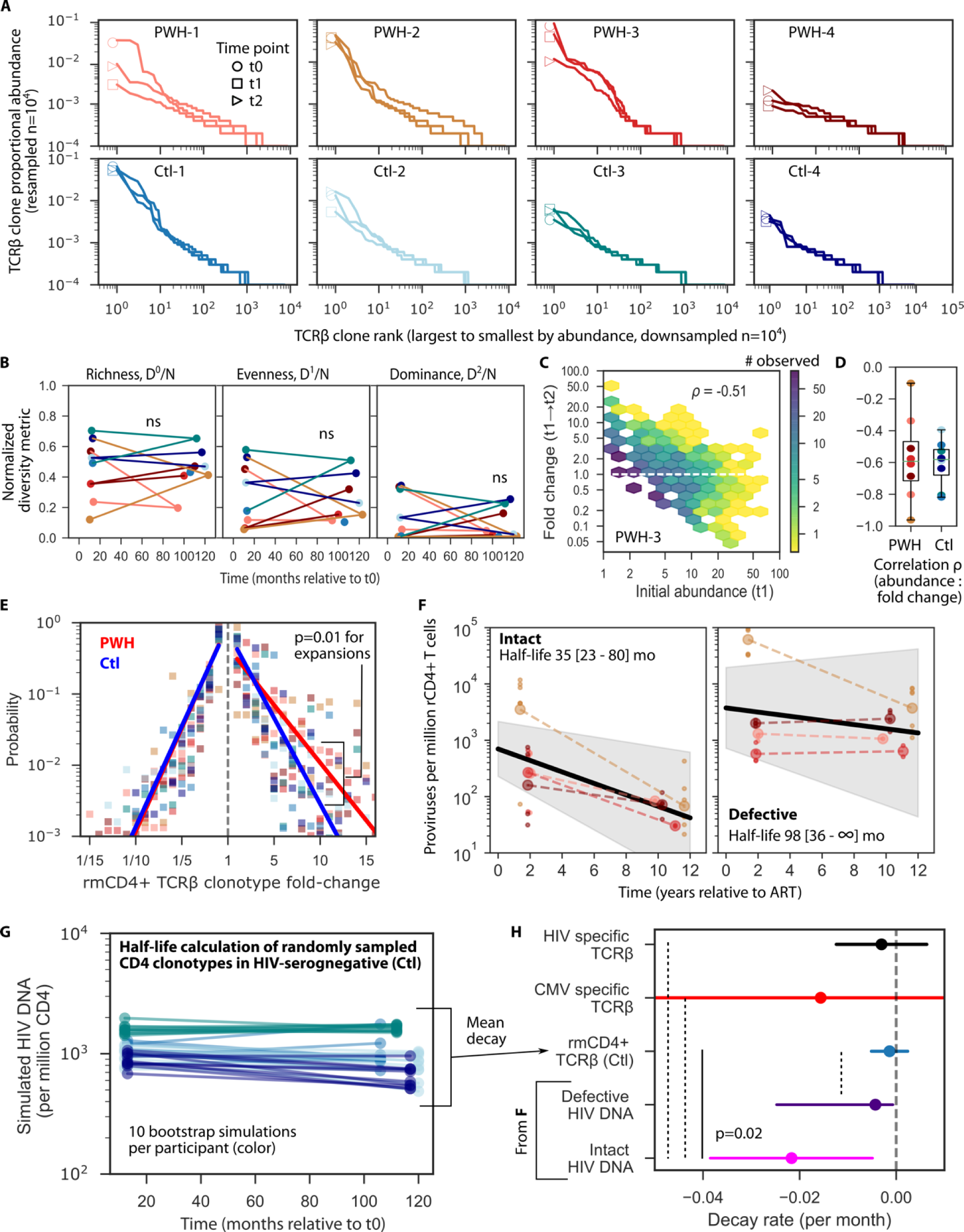
rmCD4+ TCRβ cell clones fluctuate in size and decay like defective but not intact HIV proviruses. A) TCRβ rank abundance curves from PWH and HIV-seronegative individuals sampled at 3 time points (symbol at rank=1 specifies sample time point). All data are resampled to have 10^4^ total sequences. B) Longitudinal normalized ecological metrics from persistent TCRβ. No decay was observed by Pearson correlation (ns). C) Example comparison of initial abundance at t1 vs. fold change to t2 of all rmCD4+ T cell clones that were observed in a single participant. Color bar shows how frequently a certain initial size occurred (density of observations in each hex bin). The most common clonotype is initially small and remains small (purple). Initially large clones infrequently expand, and higher observed initial abundance predicts clonal contraction (Spearman ρ=-0.5). D) Spearman correlation coefficient (as in **C**) between initial abundance and fold change for all PWH and Ctl participants. E) Across all rmCD4+ TCRβ that were observed at two time points, we calculated the probability of each TCRβ to expand or contract in abundance at a certain level. Large expansions and contractions are less and less likely to occur, roughly following an exponential distribution with fold-change. Dots indicate values from each participant and red and blue line summarizes the slope of the probability distribution for expansions and contractions in PWH and Ctl, respectively. P value (one sided Mann-Whitney) indicates significant difference between the slope of the distribution of expansions comparing PWH and Ctl cohorts. F) Observed levels of Intact and defective HIV DNA for all 4 PWH and modeled decay (population mixed effects approach to estimate decay rates) over the first 10 years of ART. Mean population decay rates are shown as black lines and 95% CI are shown as shaded intervals. Large circles, mean; small circles, replicates. G) Simulated half-life of randomly sampled of TCRβ sequences from HIV-seronegative individuals (Ctl) to mimic IPDA decay experiment (**Methods**). 10 bootstrap simulations were performed for each individual (colors). H) Observed intact and defective HIV DNA decay rates compared to the mean simulated decay rates from rmCD4+ T cells in **G** from all participants. Additionally, the average and 95% CI for HIV- and CMV-specific clonotypes between t1 and t2 is also presented. Dots are mean, lines are 95% CI for the mean. P-values indicate significant differences using Z-test. Dashed lines indicate non-significant comparisons.

### rmCD4+ clones wax and wane with time

Although rank abundance shapes are relatively constant, these distributions do not indicate whether the individual TCRβ clonotypes making up the distributions change in size (or in ranks). Therefore, we calculated fold-changes in clonotype sizes over time in PWH and Ctl participants (example in **Figure 5C**, all participant data in **Figure S10**. We found that most clonotypes which were initially small (observed singletons) did not change substantially at follow up time points, though occasionally they expanded. It was almost never observed that an initially large clone (observed abundance >30 in sample size 1e4) expanded further by the second time point. Overall, across participants a strong negative correlation was observed between initial size and fold-change in size with time (median Spearman ρ=-0.6, **Figure 5D**). That rmCD4+ T cell clones fluctuate in size or “wax and wane,” as documented for HIV proviruses^57^, despite little change in clonal structure indicates an impressive dynamic equilibrium. It appears that the identity of the clonotype inhabiting each rank in the distribution often changes even while the relative abundances between the ranks remains relatively stable. For instance, new clones may occupy the first and second ranks at a different time point, but the relative sizes of the two new clones remains very similar to the top-ranked clones from a prior or future time point.

### Expansions and contractions of rmCD4+ T cells are roughly symmetric and exponentially distributed

To further explore clonal dynamics, we quantified the frequency of expansions and contractions of rmCD4 clones, finding an approximately exponential distribution such that larger magnitude changes were less likely to occur (**Figure 5E**). rmCD4s of PWH on ART were more likely to undergo large clonal expansions when compared to HIV-seronegative participants (p=0.01) but contractions were similar across cohorts.

### Defective proviruses decay at a similar rate as general rmCD4+ T cells, both of which decay significantly slower than intact HIV proviruses

From our cohort of 4 PWH, we used the intact proviral DNA assay (IPDA) to measure intact and defective HIV proviral decay rates in the longitudinal PWH on ART and found them to be in line with results from other publications^5,10,36,79,80^: the population mean intact provirus half-life was 35 months (95% CI 23-80 months) and the population mean defective provirus half-life was 98 months (95% CI 36-infinite)^13^ (**Figure 5F**). The intact provirus decay rate is faster than the defective provirus decay rate, but it remained unclear whether either of these rates simply reflected the half-life of uninfected rmCD4s. Thus, we sought to recapitulate the decay rate experiment in our matched longitudinal cohort of HIV-seronegative people. We obtained TCRβ repertoires from total rCD4+ T cells at t1 and t2 in HIV-seronegative participants. Importantly, the total rCD4 repertoires included both memory and naïve cells, just as we measured HIV infected cells among total rCD4 cells in our IPDA experiments. We randomly sampled a subgroup of resting *memory* CD4+ T cells identified from rmCD4+ TCRβ repertoires (having the same frequency as HIV-infected cells measured by IPDA, ∼1000 per million CD4 at t1, **Figure 5F**) and then determined the change in frequency between t1 and t2 of that subgroup as a proportion of total rCD4 cells using the sum of their TCRβ abundances. We repeated this random sampling and frequency tracking for each HIV-seronegative Ctl participant 10x per participant (**Figure 5G**).

We found that the decay rate of uninfected rmCD4s was on average zero. The corresponding half-life was not statistically distinguishable from that of defective proviruses but was statistically different from that of intact proviruses, which decay over time (**Figure 5H**). This suggests that rCD4s harboring defective proviruses may generally experience similar proliferative and destructive forces in humans as HIV-uninfected rmCD4s, remaining at stable levels in circulation over ten years with little appreciable selective advantage or disadvantage due to HIV-specific or other factors. In these 4 participants, HIV-infected cells harboring intact proviruses appeared to decay faster than uninfected rmCD4 cell in the first decade of ART (p=0.02) (**Figure 5H**). Though the means were slightly different, defective provirus decay was not significantly different from that of rmCD4 clonotypes.

Finally, we calculated the decay rate of all CMV- and HIV-specific clonotypes between t1 and t2 (**Figure 5H**). These rates were not different from zero, indicating that typical HIV- and CMV-responding rmCD4s in the two individuals assayed with ViraFEST remained roughly stable over a decade of suppressive ART (**Figure 5E**). Because of small numbers and high heterogeneity, these rates have wide 95% confidence intervals.

### Simultaneous mathematical modeling of rmCD4+ TCRβ and HIV-infected cell clones

Next, we sought to design a mathematical model that was sufficient to comprehensively describe the many observations relating to both HIV-infected cell and rmCD4+ TCRβ clone dynamics presented here and elsewhere. The model is a variant merging our previous work on HIV phylodynamics^81^ and reservoir creation^70^ (**Figure 6A, Supplementary Methods**).

**Figure 6.**
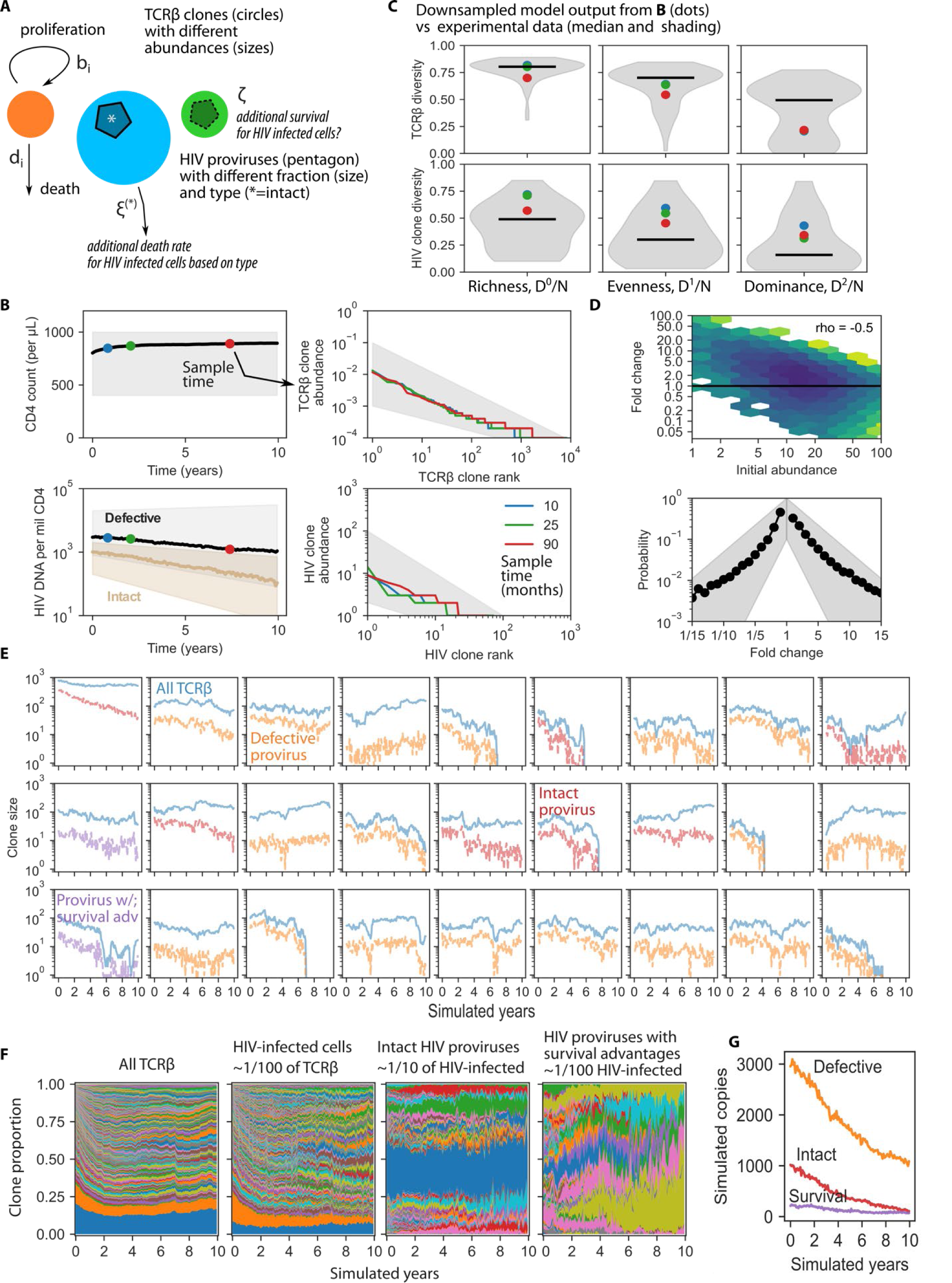
Simultaneous modeling of CD4+ TCRβ and HIV-infected clones reveals mild negative selection against HIV proviruses and allows the possibility of HIV-specific positive selection. A) Model schematic to simulate the comprehensive system linking TCRβ carriers and HIV proviral passengers during suppressive ART. TCRβ clones proliferate and die, some carry HIV (pentagon), some HIV DNA is intact (*) and some is defective (dashed line). The optimal model required an additional death rate (negative selection) on HIV-infected cells, which varies for intact and defective proviruses. B-D) Model output recapitulates ranges from experimental data (shaded regions). B) mCD4 and intact and defective HIV DNA population sizes as well as associated rank abundances at 3 time points (dots in left panel indicate sampling times corresponding to rank abundance curves of same color in right panels). CD4 counts follow general trends of experimental data and are stable while HIV DNA decays and intact decays more rapidly than defective HIV DNA. TCRβ rank abundances are static in time (overlaying one another) whereas those of HIV become more clonal over time (red high/middle rank clones are slightly larger). C) Ecological diversity metric values calculated for TCRβ and HIV provirus abundances. TCRβ are sampled at 1e4 cells and HIV provirus is sampled at 100 cells. Dots are model output from months 10, 25, and 90 as in B compared to shaded regions and medians from experimental data. D) Sampled TCRβ initial clone sizes are negatively correlated with fold change with Spearman correlation coefficient matching experimental data in **Figure 5F**. Simulated expansion/contraction distributions are symmetric and fall within boundaries (shading) of participant data in **Figure 5E. E**) 27 examples of TCRβ clonotypes (blue) with linked HIV provirus passenger levels (defective = orange, red = intact, purple = survival advantage) illustrate representative fluctuations in clone sizes and illustrate that TCRβ clones can have a heterogeneous fraction of their associated cells that carry HIV proviruses. F) Proportional clone sizes for TCRβ, total HIV infected cells, intact HIV proviral clones, and survival advantaged HIV clones. G) Total levels on non-log scale of each proviral type over 10 years of simulation time.

Briefly, rmCD4+ TCRβ clones *T*_*i*_ are distributed into *T*(0) =10^6^ rmCD4+ T cells such that ∑_*i*_ *T*_*i*_ = *T*(0). We set the fraction of cells carrying HIV DNA (*H*(0) = 3000 per million CD4s) and intact (*H*^∗^(0) = 800 per million CD4s) proviruses using population means from our data (**Figure 5F**). The size of each rmCD4+ T cell clonotype is initialized by drawing from a distribution. Then, TCRβ clonotypes are given proliferation (*b*_*i*_) and death rates (*d*_*i*_), also drawn from a distribution. Thus, the size of each TCRβ clone on average follows the differential equation

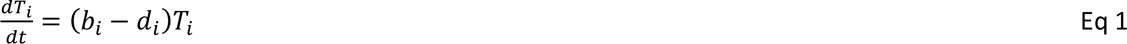

Stability is ensured on average by setting *b*_*i*_ = *d*_*i*_ for each clone. We simulate the model as a discretized stochastic system (**Methods**) such that clones expand and contract sporadically, those with higher birth and death rates do so more rapidly, and any clone can go extinct and be removed if its abundance reaches zero.

### Sequential inclusion of model features to recapitulate experimental data

We sought a model that could output six key data features/observations simultaneously: 1) stability of rmCD4+ T cell population sizes over 10 years (**Supplementary Figure 1**), 2) decline of intact and slight or no decline of defective proviruses, matching prior published observations of these half-lives^8–14^ (**Figure 5F**), 3) fluctuations of clone sizes with waxing and waning of both rmCD4 cell and HIV-infected cell abundances as we (**Figure 5C&D**) and others^57,82^ have observed, 4) a roughly exponential and symmetric distribution of rmCD4+ TCRβ expansions and contractions (**Figure 5E**), 5) a power law rank abundance (and associated ecological metrics) of rmCD4+ TCRβ clonotypes that is stable over time (**Figure 5A&B**), and 6) a highly downsampled power law rank abundance of HIV clonotypes that becomes more clonal over time and matches metrics from participant data.

In **Supplementary Data 1**, we provide simulation output from all models that were tested (annotated as **Sim1** through **Sim6**), the sequential complexity that was added to each model, and any experimental observations each model could not recapitulate. For instance, we first assumed initial conditions and proliferation/death rates were drawn from uniform distributions. This model did not achieve agreement with TCRβ and HIV proviral clonality (**Sim1**). Instead, we found that initializing clonotypes with a power-law distribution^83^ *T*_*x*_(*t* = 0)∼*x*^−*z*^ where *z*=0.8 could achieve fit to observed TCRβ clonality and rank abundances (**Sim1**). A key requirement for the model to achieve the simultaneously stable rmCD4 levels and rank abundance while also admitting waxing and waning was that individual clonotypes could not have the same birth and death rate for the entire simulation (10 years). We found redrawing (or “shuffling”) these rates every *s*=100 days (**Sim4**) was sufficient to achieve qualitatively reasonable behaviors, but the exact value *s* could not be precisely identified from the present data. In addition, an exponential distribution for proliferation and death rates *b*_*x*_ = *d*_*x*_∼*α*_*T*_exp(−*x*) parameterized by the average proliferation rate of central memory CD4+ T cells^24,84^, or *α*_*T*_=0.01 per day, was preferred over a uniform distribution with the same mean to quantitatively match observed ecology and expansions and contractions of clones over time (**Sim6**).

### Simulations of the optimal model show agreement with HIV and CD4+ TCRβ clonality and clonal dynamics

The optimal model (**Sim6**) matches several types of observational data (**Figure 6B&C**). First, numbers of rmCD4s fall in boundaries (shaded areas, data from **Figure S1**) of general CD4+ T cell count trends from PWH on ART (**Figure 6B**). Second, rmCD4 clonotype rank abundances sampled at n=1e4 from the model are power-law distributed, stable over time, and fall in boundaries of participant data (shaded areas, data from **Figure 1C**). Third, intact and defective HIV DNA levels decay with half-lives matching population data (shaded areas, data from **Figure 5F**). HIV rank abundances sampled at n=100 at 3 time points appear like undersampled power law distributions, fall within boundaries of PWH data (shaded areas, data from **Figure 1D**) and become more clonal over time. Third, using downsampled data and calculating ecological metrics as in our analysis of experimental data, model output simultaneously matches clonality of both TCRβ and HIV proviruses (shaded areas, data from **Figures 1&2**) (**Figure 6C**). Fourth, the model matches observed TCRβ clone dynamics, outputting both a correlation between initial clone size and fold-change (Spearman ρ=-0.5), in excellent agreement with experimental observations (data from **Figure 5C**) (**Figure 6D**, top), and a symmetric distribution of expansion/contraction sizes that agrees with distributions from experimental data (shaded areas, data from **Figure 5E**) (**Figure 6D**, bottom).

### Modeling reveals mild negative selection on intact and some defective proviruses

To recapitulate intact and defective proviral decay rates while maintaining all rmCD4 clonality dynamics and ecology, we required cells carrying intact (and to a lesser degree, defective) proviruses to be modulated with an additional death rate that hypothetically represents some process of viral reactivation and/or immune recognition followed by immune clearance. Each clonotype was assumed to have an additional death rate *ξ*\*=6.5e-4 (intact) and *ξ*=2.3e-4 (defective) per day depending on whether the provirus is intact vs defective^61,62^.

However, the absolute magnitude of these additional rates in comparison to the natural death rates of CD4+ T cells is meaningful. The average proliferation and death rates in the model (⟨*b*_*i*_⟩ and ⟨*d*_*i*_⟩) were set to those from central memory T cells, which turn over ∼3.7 times per year, or 0.01 per day^24^. This means that in our simulation, the fraction of proviral clearance that is due to HIV-specific selection is approximately *ξ*\*/(*ξ*\* + ⟨*d*_*i*_⟩)=6% for intact proviruses and 2% for defective proviruses, where ⟨*d*_*i*_⟩=0.01 per day, the average death rate in the model. Natural clearance is balanced by birth for CD4+ T cells, but over years on ART, the additional death rates on HIV-infected cells amount to a slowly decaying pool of proviruses.

One final observation on the optimal model structure is that increases in HIV clonality without commensal changes in TCRβ clonality did not require individual clones that were permanently skewed toward clonal expansion --as they might be if HIV integration site or stable pro-survival signals drove proliferation^27,46^.

### Slight survival advantages for rare intact and defective proviruses could be incorporated into the model without changing observed dynamics over 10 years of ART

It has been observed that certain infected cells may have some survival advantages compared to uninfected cells based on integration site locations or other mechanisms^44,45,47–50,85^. As a sensitivity analysis, we therefore added a positive “survival advantage” rate *ζ*_*i*_=1e-5 per day for a small fraction of HIV-infected clones *f*_*ζ*_=1/100. This rate was not dependent on whether the HIV provirus was intact or defective. The inclusion of this mechanism at this magnitude did not substantially change our model output comparison with experimental data. However, setting *f*_*ζ*_=1/10 or *ζ*_*i*_=1e-4 did disturb reservoir dynamics and resulted in overly clonal/stable reservoirs.

In conclusion, we iteratively generated a model (**Figure 6A**) that matches all desired qualitative and quantitative observations. The form of this ultimate model illustrates the forces on a single HIV infected clone as a balance of proliferation and death rates of the CD4+ T cell clonotype into which it is integrated – governed by Equation 1, less some additional death due to immune selection (or other mechanism) that acts differently on intact *H*\* vs defective *H*^()^ HIV proviruses, and potentially plus some survival advantage related to integration site or other pro-survival mechanisms:

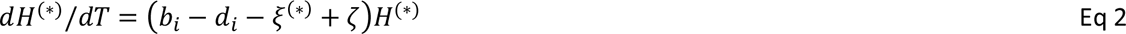

### Model projections of individual clone dynamics

Using this final model, we could further illuminate the predicted underlying clone dynamics that generate outputs. For instance, as observed previously in PWH on ART^41^, the model also predicts that HIV-infected CD4 clonotypes are heterogeneous in the proportion of clonotype cells carrying given proviruses: some clonotypes have very few of their cells infected, whereas other clonotypes have a majority of their cells infected --driven in the model by the stochastic trajectory, the size of the TCRβ clonotype when it was initially infected, and the type of provirus carried (**Figure 6E**)^15^. HIV-infected cells carrying intact proviruses tend to decrease in fraction within individual TCRβ clones (**Figure 6E**, examples in red), whereas the rare HIV-infected cells carrying proviruses with survival advantages remain stable or grow (**Figure 6E**, examples in purple). The clonal proportions of each type of cell or provirus are relatively stable over time by eye, but HIV-infected cells are slightly more clonally dominated, and the rarer intact/survival advantaged clonotypes are still more clonal (**Figure 6F**). Finally, given the rarity of proviruses with survival advantages required to match the experimental data, we project they do not make up a large proportion of HIV infected cells early on ART. However, after 10 years of simulations, levels of these proviruses begin to approximate intact provirus levels (**Figure 6F**). This projection may reconcile data from PWH on ART >15 years suggesting intact proviral decay may slow or stop entirely (potentially even transitioning to growth)^10–12,36,80^.

## Discussion

In this study, we investigated whether the natural physiology of mCD4+ T cells can entirely explain the clonality and decay dynamics of the HIV reservoir in PWH on ART, or whether positive or negative selective forces acting on HIV-infected cells are also required to explain these observations. We focused on three key observations regarding HIV-infected CD4s in PWH on ART: 1) the high degree of reservoir clonality, 2) the increase in HIV-infected cell clonality over time on ART, and 3) the differential decay of CD4s harboring intact vs defective proviruses over time on ART. We found that the natural biology of mCD4+ T cells dovetails with the first observation but cannot explain the second and third observations, implying that selective advantages or disadvantages due to factors specific to HIV-infected cells contribute to the clonal kinetics of the reservoir. Moreover, we report that the shallow provirus sampling depths common in observational studies prevent accurate comparisons of the full clonality distributions between HIV-infected cells and rmCD4+ T cells. Without a full characterization of the ‘tail’ or ‘second slope’ of provirus clonality distributions, we cannot know to what degree forces like antigen stimulation and homeostatic proliferation maintain the reservoir in its totality in PWH on ART. Finally, we construct a mathematical model of HIV-infected cell and reservoir dynamics in PWH on ART through iterative testing and comparison to observational data. In this model, HIV-specific negative selective forces are mild in scope, responsible for approximately 2-6% of cell death events in mCD4s harboring defective and intact HIV proviruses. This model allows for HIV-specific pro-survival selective forces, such as integration into transcriptionally silent genomic regions or upregulation of pro-survival cellular programs, at a very low level (≤1% of HIV-infected cells) at ART initiation. Over the years on ART, the proportion of these survival-advantaged HIV-infected cells among all HIV-infected cells grows, perhaps explaining why the decay of the reservoir slows over time on ART^10–12,36,80^.

Regarding the first observation, many groups have observed the high degree of clonality in the reservoir and in HIV-infected cells (intact plus defective) in PWH on ART, reviewed previously^86,87^. Some groups have demonstrated that HIV-infected cell clonality is greater than that of memory CD4s from the same sample^39,85^ but have not controlled for new diversity entering the mCD4 compartment after ART initiation. This is important because generally no new HIV-infected clonotypes are generated in people on suppressive ART due to absence of viral replication^34,68,69,88^, whereas over many years, naïve CD4s encounter antigen and proliferate, with some surviving to join the long-lived mCD4 pool^83,89^. We attempt to control for that in our investigations by examining ‘persisting’ rmCD4s – those memory clonotypes that were present close to the start of ART and persist 2 and 11 years after ART initiation. We demonstrate here that HIV-infected cell clonality in PWH on ART is greater than that of persisting rmCD4s when clonal richness (number of clones per normalized sample size) and evenness are quantified.

However, we observe that measures of clonal dominance are similar between persisting rmCD4s and HIV-infected cells. Additionally, we find that measures of clonal dominance in rmCD4s, whether quantified by normalized Hill number D^2^ or a novel two-slope rank-abundance model, strongly associate with CMV serostatus, and we hypothesize that dominant clones in the reservoir and among rmCD4s become dominant due to proliferation after recent or recurrent antigen exposure. In the two longitudinal PWH available for present-day sampling, we identified TCRβ sequences of mCD4s that proliferated significantly *ex vivo* after exposure to HIV antigens and after exposure to CMV antigens. We then showed that the clonality of these antigen-responsive clones was similar to that of HIV-infected cells across all measures of clonality -richness, evenness, and clonal dominance. This implies that natural forces acting on mCD4s, in this case antigen responsiveness, can account for the clonality of the observed HIV reservoir.

We document an important caveat to this conclusion. While we demonstrate that our sampling depths of rmCD4+ T cells reach clonal richness saturation, we demonstrate that HIV provirus sequencing depths of up to 100 sequences per time point do not appear close to richness saturation (**Figure 1C**). This means that provirus sequencing depths of greater than 100, and probably greater than 1000 are needed to approximate the richness of HIV-infected clones in a given sample and to assess the clonality of non-dominant HIV-infected clonotypes. However, the labor-intensive nature of provirus sequencing and the rarity of HIV-infected cells in single cell RNA sequencing approaches make it prohibitively time-consuming or expensive to reach HIV provirus sampling depths much beyond 100. One group has directly compared HIV integration site datasets – which do not distinguish intact from defective proviruses -on the order of 1000+ per sample to mCD4 TCRβ repertoires and found that the largest HIV-infected clones are more stable than randomly-chosen mCD4+ T cell clones of the same size^35^. One plausible explanation for this is that the largest HIV-infected clones are biologically more similar to the largest randomly-chosen mCD4+ T cell clones instead of the mid-sized clones with which they were compared. We show here that the largest HIV-infected cells and the largest mCD4+ T cells can be modeled with the same power-law slope, and we think it possible that both types of clones are responding to recent or recurrent antigen. Our work reveals that undersampling of reservoir datasets leads to characterization of the most highly expanded, or clonally dominant, HIV-infected cells, whose dominance is likely driven by exposure to antigen. Thus the HIV cure field may be biased towards results implying that antigen-driven proliferation provides the dominant stimulus for the persistence of the HIV latent reservoir. We show that the clonality and the longitudinal dynamics of non-dominant HIV-infected clones – the ‘tail’ or the second slope of the clonality distribution -have not yet been characterized (**Figure 3F**), although non-dominant clones probably make up the majority of HIV-infected cells in humans^34,35,38^. Because of this gap in knowledge, it remains unknown whether homeostatic proliferation, less frequent or intense antigen exposure, or other forces underlie the persistence of the reservoir in non-dominant clones. In order to understand the persistence of the HIV reservoir in its entirety in PWH on ART, we need more deeply sampled reservoir datasets.

Regarding the second observation, we and others have demonstrated that the clonality of HIV-infected cells increases with time on ART^13,14,27,37,90^. Our previous work modeling this change in clonality suggested that after a decade on ART, over half of an individual’s remaining proviruses are members of the top 100-1000 most-highly expanded infected clones^13^. Here we demonstrate that mostly-uninfected persisting rmCD4s from PWH on ART and uninfected rmCD4s from HIV-seronegative people do not increase in clonality with time using any of the four measures of clonality we examine. Additionally, we did not observe increases in clonality over ten years on ART of the antigen-specific clonotypes we examined, although our observations of antigen-specific cells were limited by numbers of people available for current-day sampling and should be repeated with more participants. In addition, proliferation *ex vivo* upon stimulation with high peptide/antigen density may capture only some of the clones responding to a certain pathogen *in vivo*. Taken together, HIV-specific forces seem to be required to explain observations of increasing HIV-infected cell clonality over time.

Regarding the third observation, many studies have now documented a decay in rCD4+ T cells harboring intact HIV proviruses in the first 7-10 years on ART and a relatively stability, or at least an extremely slow rate of decay, in rCD4+ T cells harboring defective HIV proviruses in PWH on ART^8,10,13,36,91^. We demonstrate here that decay rates of defective proviruses are similar to decay rates of mostly-uninfected rmCD4s in PWH on ART, HIV-responding rmCD4s, and uninfected rmCD4s in HIV-seronegative people. Importantly, we show that clearance of intact proviruses would not be predicted by HIV-independent rmCD4+ T cell forces alone^59,64^. Overall, this suggests that the presence of intact provirus in mCD4s is associated with enhanced death rates, at least in the first decade on ART. Our observational data still allows the possibility that defectiveness is a continuum and that certain defective clones experience more or less negative selection.

We compared normalized diversity metrics across our carefully matched longitudinal cohorts and found that rmCD4+ T cell diversity metrics did not distinguish participants with suppressed HIV infection from those who were HIV-seronegative and matched on age, sex, race, and CMV serostatus. One difference we observed is that clonal expansions of rmCD4 cells over time was slightly more common in PWH on ART than HIV-seronegative people, but we could not tell whether this was due to CMV, which is associated with clonal expansion over time, or other factors because all but one longitudinal participant had CMV. Beyond HIV, the longitudinal nature of our study admitted some novel properties of rmCD4+ T cell repertoires. First, although repertoires had distinct rank-abundances shapes for each individual, rank-abundances were remarkably stable within participants – including PWH on ART (**Figure 1 & 5**). Although aging is known to modulate T cell diversity^92,93^, the repertoire clonality among these adults did not substantially change over a decade of observation. Second, we observed that individual clonotypes are dynamic with constant shifts in the dominant clonotypes. Large clones observed at one time point were much more likely to contract than expand at a later time point (**Figure 5C**). With respect to HIV reservoirs, dynamic TCRβ expansions and contractions (waxing and waning) serve as a potential mechanistic basis for observed HIV clone waxing and waning^57^. Third, we observed in HIV-seronegative people that, in contrast to intact HIV DNA levels, the frequency of a randomly selected group of rmCD4 clones among total resting CD4+ T cells (memory plus naïve) does not change in abundance over a decade of observation, a powerful example of the maintenance of immune memory in humans.

Our introduction of the double power law model of rmCD4+ T cell clonality illuminated that a model that separated TCRβs into two distinct clonality categories was associated strongly with CMV serostatus, hinting that (as observed previously using other approaches^73^) large TCRβ clones could be responding to CMV. When we directly tested which rmCD4 clones retained the capacity to proliferate to CMV antigen, we were able to identify only one of the largest clones from years ago as currently CMV-responsive, while many smaller clones were identified as currently CMV-responsive --similar to other groups’ findings^85^. This may be due to assay insensitivity (wherein most large clones respond to CMV but our assay missed them), or because large clones are responding to other antigens (including bacteria^94^), or because the antigen-specific large clones identified at years 2 or 11 after ART initiation are now functionally exhausted at year 16 or 19.

Our mechanistic model grows from prior work on T cell dynamics and distributions^83,95–99^ and proliferative HIV clone dynamics (modeled similarly to our approach^100^ and in more classical viral dynamics models^101,102^) into what we believe is the most rigorously data-validated model to describe the linked system of uninfected and infected CD4+ T cells in PWH on ART^74^. It achieves our goal of matching many observed features of HIV and CD4+ TCRβ clonal and population size dynamics simultaneously. It infers a slight but essential force of negative selection on HIV-infected cells that we estimate accounts for ∼2% of defective and ∼6% of intact HIV provirus clearance during ART^48^. Additionally, it supports the possibility of rare (<1/100) proviruses that attain survival advantages based on integration site, viral genetic features, or upregulated pro-survival cellular programs, phenomena now observed by many groups^26,27,37,42–45,47–50,85^ that could explain reservoir stability or expansion after decades of ART. Still more or different data would be needed to estimate this population’s size and dynamics precisely in our framework. Of course, the model naturally oversimplifies the system. Among many approximations, clone proliferation is stochastic but does follow programmed expansions and contractions^103^, and we also institute an artificial reshuffling of clone proliferation rates. For now, we do not directly connect cellular proliferation to viral reactivation, as it is not yet clear these are consistently linked^104–106^.

There are important limitations in this study. We sequenced near-full length (nfl) proviruses, QVOA-outgrowth HIV *env*, and TCRβ, and each method has different biases in approximating clonality, so their comparison is not perfect. Nfl clonality is strong evidence of true clonality^108^, especially for defective proviruses, while HIV *env* sequencing is likely biased towards higher clonality despite being the most genetically diverse region of the HIV genome. TCRαβ repertoires would be ideal for T cell clonality determination, though its expense and depth limit its use presently. A particular caveat is that we sequenced resting CD4+ T cells here, as activated CD4+ T cells may be actively dividing and short-lived^21,24,109^. We might have missed important clonal dynamics in lymphoid associated and tissue-resident CD4, where central memory cells may be marginated until they are activated to become effector memory cells and enter the circulation. Another consideration is that more than one HIV provirus may be integrated into the same rmCD4+ T cell or clonal population. However, multiple proviruses are not often found in the same cell^110^, and if multiple proviruses were harbored by the same TCR clone this would only add ballast to the argument that HIV dynamics are driven by the largest clonotypes. Although the size of our cohort was limited by the time and expense of the assays, strengths of our study include the integration of large, published data sets and our novel matched longitudinal cohorts, in which PWH with documented ART adherence over 10+ years were age/race/sex matched with HIV-seronegative people.

In the end, our data and modeling promote a picture of powerful rmCD4+ T cell homeostasis between clonal birth and death, with dynamic clonal reshuffling underlying a remarkably stable overall clonality structure within each individual over the course of a decade in mid-life^107^. HIV proviruses are similarly clonal to the most clonally-dominant mCD4+ T cell clones, which are likely antigen specific, but deeper sampling is needed to understand the clonality and dynamics of the less clonally-dominant reservoir. Additional forces of negative selection on intact and some defective proviruses are required; other competing pro-survival mechanisms for proviruses may exist and addition of this modeling term reconciles HIV reservoir stability or even expansion after a decade of ART. In conclusion, the dynamics and clonality of HIV proviruses appear to be predominantly, but not completely, driven by mCD4 T cell kinetics.

## Data Availability

All data produced in the present work will be freely available at the time of peer reviewed publication.

https://clients.adaptivebiotech.com/pub/deneuter-2018-cmvserostatus

## Acknowledgements

We thank all the participants who donated their time and samples. We are grateful for discussions of this work with Alison Hill, Andrew Timmons, and Nicole Walch.

Samples and data in this manuscript were collected by the Multicenter AIDS Cohort Study (MACS) and the Women’s Interagency HIV Study (WIHS), now the MACS/WIHS Combined Cohort Study (MWCCS). MWCCS (Principal Investigators): Baltimore CRS (Todd Brown and Joseph Margolick), U01-HL146201; Data Analysis and Coordination Center (Gypsyamber D’Souza, Stephen Gange and Elizabeth Topper), U01-HL146193; Northern California CRS (Bradley Aouizerat, Jennifer Price, and Phyllis Tien), U01-HL146242. The MWCCS is funded primarily by the National Heart, Lung, and Blood Institute (NHLBI), with additional co-funding from the *Eunice Kennedy Shriver* National Institute Of Child Health & Human Development (NICHD), National Institute On Aging (NIA), National Institute Of Dental & Craniofacial Research (NIDCR), National Institute Of Allergy And Infectious Diseases (NIAID), National Institute Of Neurological Disorders And Stroke (NINDS), National Institute Of Mental Health (NIMH), National Institute On Drug Abuse (NIDA), National Institute Of Nursing Research (NINR), National Cancer Institute (NCI), National Institute on Alcohol Abuse and Alcoholism (NIAAA), National Institute on Deafness and Other Communication Disorders (NIDCD), National Institute of Diabetes and Digestive and Kidney Diseases (NIDDK), National Institute on Minority Health and Health Disparities (NIMHD), and in coordination and alignment with the research priorities of the National Institutes of Health, Office of AIDS Research (OAR). MWCCS data collection is also supported by UL1-TR000004 (UCSF CTSA) and UL1-TR003098 (JHU ICTR). The authors gratefully acknowledge the contributions of the study participants and dedication of the staff at the MWCCS sites. Additional data and samples were collected from the Hospital of the University of Pennsylvania and from Philadelphia FIGHT.

The following reagents were obtained through the NIH HIV Reagent Program, Division of AIDS, NIAID, NIH: Peptide Pool, Human Cytomegalovirus (HCMV) pp65, ARP-11549, contributed by DAIDS/NIAID, Peptide Pool, Human Immunodeficiency Virus Type 1 Subtype B (Consensus) *gag* Region, ARP-12425, contributed by DAIDS, NIAID, Peptide Pool, Human Immunodeficiency Virus Type 1 Subtype B (Consensus) *env* Region, ARP-12540, contributed by DAIDS, NIAID, Peptide Pool, Human Immunodeficiency Virus Type 1 Subtype B (Consensus) *pol* Region, ARP-12438, contributed by DAIDS/NIAID, Peptide Pool, Human Immunodeficiency Virus Type 1 Subtype B (Consensus) *nef* Region, ARP-12545, contributed by DAIDS/NIAID, T-20 (N-acetylated derivative; Enfuvirtide acetate salt), ARP-12732, contributed by DAIDS/NIAID (produced by RayBiotech, Inc.).

This research was funded in part by awards (AA, KS, DBR) from the Johns Hopkins University Center for AIDS Research and the University of Washington/Fred Hutch Center for AIDS Research, NIH funded programs (P30AI094189, P30AI02775), which are supported by the following NIH Co-Funding and Participating Institutes and Centers: NIAID, NCI, NICHD, NHLBI, NIDA, NIA, NIGMS, NIDDK, NIMHD. This research was also funded in part by the NIH (grants K25AI155224 -DBR, R01AI150500 -JTS, K08AI143391 -AA, K23AI157875 -MJP, UM1AI126620 -LJM, P30AI094189 -FRS, UM1 AI164566 -FRS, U01 AI177211 –FRS, DP5OD031834 -FRS), the W.W. Smith Charitable Trust (AA), the Pearl M. Stetler Foundation (AA), and the Robert I. Jacobs Fund of The Philadelphia Foundation (LJM). LJM is supported by the Herbert Kean, M.D., Family Professorship. RS is a Howard Hughes Medical Institute (HHMI) Fellow. The content is solely the responsibility of the authors and does not necessarily represent the official views of the NIH.

## Author contributions

Conceptualization, AARA and DBR; formal analysis, DBR and AARA; investigation, AARA, DBR, DNR, AR, HZ, LZ, KNS; resources, AARA, DBR, JV, JDS, RFS, LJM, MJP, SGD, RH, LHR, SJG, NRR, PCT, JBM; writing-original draft, AARA and DBR; writing-review&editing, DBR, AARA, FRS, NRR, PCT, SGD, JTS, RFS; funding acquisition, AARA and DBR.

## Declaration of interests

MJP reports participation on a data safety and monitoring board for American Gene Technologies.

## Code availability

All code to generate all figures and perform simulations will be freely available from https://github.com/dbrvs/TCR-HIV at the time of peer-reviewed publication. Files are jupyter Python notebooks and make use of the *numpy*, *scipy*, and *seaborn* packages for modeling and data analysis.

## Study participants and clonality data

Participant data are collected in **Table 1** and **Table S1**. For this manuscript, we use the naming convention: PWH22 = PWH-1, PWH548 = PWH-2, PWH746 = PWH-3, and PWH583 = PWH-4. The linkage between participant ID and any identifying information is not known to anyone outside the research groups.

### Longitudinally sampled PWH on ART

Four research participants were selected for exceptional viral suppression over 10+ years and longitudinal samples were obtained. PWH-1 had donated samples to the Johns Hopkins HIV Clinical Cohort and research labs at Johns Hopkins, and three participants (PWH-2, −3, and −4) to the SCOPE and OPTIONS cohorts at UCSF. Sample timing, viral load, and absolute CD4+ T cell count data for these four participants are depicted in **Figure S1**. Time point 0 (t0) was a mean of 9.6 months after ART initiation (range 6.3 – 11.0 months). Time point 1 (t1) was a mean of 21.3 months after ART initiation (range 16.6 – 24.0 months). Time point 2 (t2) was a mean of 128.4 months after ART initiation (range 117.6 – 140.2 months). Time point 3 (t3) was obtained for the ViraFEST assay in the two participants willing and available to return for a dedicated research blood draw and was obtained at 189 months after ART initiation in PWH-1 and at 230 months after ART initiation in PWH-4.

Each participant maintained undetectable viral loads — checked at least every 6 months but typically more often — beginning shortly after starting ART through t2, with very few exceptions as follows (**Figure S1**): PWH-2, between t1 and t2 (10.3 years), 2 viral loads were 151 and 93 copies/mL; PWH-3, between t1 and t2 (8.4 years), 1 viral load was 42 copies/mL. Also, there were 6 instances within the >40 participant-years of follow-up in which viral loads were measured more than 6 months apart: PWH-1, once at 9 months apart; PWH-2, once at 7 months apart, once at 12 months apart, and 3 times at 9 months apart. PWH-1, −2 and −4 had negative Hepatitis C virus (HCV) IgG tests after t2, and participant PWH-3 had a negative HCV IgG test after t1 and 3.5 years before t2, with no further HCV testing available.

### Longitudinally sampled HIV-seronegative people

Cryopreserved longitudinal samples from one female participant of the WIHS and three male participants from the MACS were obtained^51^. These participants were confirmed to be HIV-seronegative at all sampled timepoints. Participants and samples were matched one-to-one on age, race, sex, and time between samples to the longitudinally sampled PWH on ART. These participants were all HCV seronegative.

### Cross-sectionally sampled PWH on ART

*Env* sequences from limiting dilution quantitative viral outgrowth assays from seven participants in a previously published study were obtained^52^. Only *env* sequences from wells that were not treated with autologous IgG were used in this analysis. The HCV serostatus of this cohort is unknown.

### Cross-sectionally sampled HIV-seronegative people

Single-timepoint memory CD4+ T cell receptor β chain repertoires from a generally healthy cohort of 33 Belgian individuals were obtained from a public database (De Neuter^53^). These individuals were all HIV negative at the time of sampling (Dr Benson Ogunjimi, personal communication). It was not possible to match these individuals on demographic characteristics to the cross-sectionally sampled PWH on ART. The HCV serostatus of this cohort is unknown.

## Study approval

The studies from which samples were obtained were approved by the UCSF Committee on Human Research, the Johns Hopkins University School of Medicine Institutional Review Board (IRB), and the University of Pennsylvania IRB. All participants provided written informed consent before enrollment.

## HIV provirus sequencing

Near-full length HIV provirus sequences from resting CD4+ T cells from the four longitudinally sampled PWH on ART were obtained from a previously published study^13^. *env* sequences from limiting dilution quantitative viral outgrowth assays of resting CD4+ T cells from seven participants in a previously published study were obtained^52^. Only *env* sequences from wells that were not treated with autologous IgG were used in this analysis.

## Flow sorting of resting memory CD4+ T cells

Viably frozen PBMCs were quick thawed at 37°C, rinsed, centrifuged (5 minutes, 500 g), resuspended, and rested overnight in an incubator. Cells were incubated with Human BD Fc Block (BD Pharmingen) at room temperature for 10 minutes. Cells were stained, followed by a 30-minute incubation on ice, with an APC-labeled antibody against CD3 (BioLegend; clone UCHT1), PE-Cy7–labeled antibody against CD4 (BioLegend; clone RPA-T4), BV421-labeled antibody against CD45RA (BioLegend; clone HI100), PE-labeled antibody against CCR7 (BioLegend; clone G043H7), FITC-labeled antibodies against CD69 (BioLegend; clone FN50) and HLA-DR (BioLegend; clone L243), and PE-Cy5–labeled antibodies against CD14 (Thermo Fisher Scientific; clone 61D3), CD16 (BioLegend; clone 3G8), CD20 (BioLegend; clone 2H7), and CD8a (BioLegend; clone RPA-T8). Dead cells were excluded using propidium iodide. Cells stained with single fluorophore-labeled antibodies were used to set sorting gates. A representative gating strategy and sorting logic is provided in **Figure S2**. CD45RA expression was used to distinguish naïve-like (Na) and terminally differentiated cells from memory cells. Central memory (Tcm) cells were distinguished from effector memory (Tem) cells by the expression of CCR7. Cells were sorted using the Beckman Coulter MoFlo Legacy cell sorter.

## Isolation of total resting CD4+ T cells

To obtain total (memory and naïve) resting CD4+ T cell TCRβ repertoires for the denominator of frequency measurements of random groupings of rmCD4 cells in HIV-seronegative participants, viably frozen PBMCs were quick-thawed at 37°C, rinsed, centrifuged (5 minutes, 500 g), resuspended, and rested overnight in an incubator. Total CD4+ T cells were purified using an immunomagnetic negative selection kit according to the manufacturer’s instructions (EasySep^TM^ Human CD4+ T Cell Isolation Kit, STEMCELL Technologies). Immediately afterwards, resting CD4+ T cells (CD4+CD69-CD25-HLA-DR-) were enriched by a second negative depletion (biotin-conjugated anti-CD25, anti-biotin MicroBeads, CD69 MicroBead Kit II, and anti-HLA-DR MicroBeads, Miltenyi Biotec).

## TCRβ repertoire sequencing

Genomic DNA was isolated (QIAamp DNA Mini or Micro kit, Qiagen, or Quick-DNA Miniprep, Zymo Research) from sorted cells, quantified (Qubit 3.0 or 4 fluorometer, dsDNA quantitation, high sensitivity or broad range assays, ThermoFisher Scientific, or by NanoDrop 2000 spectrophotometer, ThermoFisher Scientific) and diluted in Tris-acetate-EDTA to a concentration of 10-50 ng/μL (for a total of up to 1-3 μg per sample). TCRβ sequencing data were generated using the immunoSEQ hsTCRβ assay, version 4, deep or ultradeep mode (Adaptive Biotechnologies). TCRβ sequences for the ViraFEST assay were generated using the AmpliSeq for Illumina TCR beta-SR panel (Illumina).

## Identification of HIV-specific and CMV-specific CD4+ T cells: ViraFEST assay

The viral functional expansion of specific T cells (ViraFEST) assay identifies TCRβ clonotypes that proliferate in response to viral antigen^75,111,112^. Once antigen-specific TCRβ DNA sequences were identified by ViraFEST, their corresponding amino acid sequences were identified as antigen-specific within the same participant. When quantifying abundances of antigen-specific TCRβs for a participant, any TCRβ DNA sequence that yielded an antigen-specific amino acid sequence for that participant was considered antigen-specific. As outlined in **Figure 4A**, CD8-depleted PBMCs from a fresh blood draw are stimulated with either HIV or CMV viral antigen in the presence of antiretroviral drug to induce antigen-specific expansion followed by TCRβ repertoire sequencing of wells to identify clonally expanded, antigen-responsive TCRβ sequences. At time point 3 (t3), over 80 mL of peripheral blood was obtained from PWH-1 at 189 months and PWH-4 at 230 months after ART initiation using an acid-citrate-dextrose or ethylenediaminetetraacetic acid (EDTA) anticoagulant and PBMCs were isolated using a Ficoll density gradient as previously described^113^. CD8 T cells were depleted from PBMCs (CD8 Dynabeads, ThermoFisher Scientific). 2 million CD8-depleted PBMCs were plated in 2mL culture medium (IMDM, 5% human serum, 50 µg/mL gentamicin) containing 10 IU/mL IL-2 and 10µM enfuvirtide. To each well one of the following was added: (1) either 5 µg for PWH-4 or 10 µg for PWH-1 peptide/mL HIV-1 consensus B *gag*, *pol*, *env*, and *nef* overlapping peptide pools (NIH HIV Reagent Program), (2) 10 µg peptide/mL CMV pp65 protein overlapping peptide pool (NIH HIV Reagent Program) and 3 µg/mL gradient-purified inactivated CMV (Virusys), (3) 0.5% sterile DMSO by volume as a negative control, or (4) anti-CD3/CD28 antibody-bound beads in a 1:1 bead:cell ratio as a positive control (Human T-Activator CD3/CD28 Dynabeads, ThermoFisher Scientific) and incubated at 37° C. Each assay condition was performed in triplicate. On days 3 and 7, half the media was replaced with fresh culture media containing IL-2 and enfuvirtide. Wells were split on day 7 if needed. On day 10, cells were harvested and CD4+ T cells were isolated using the EasySep CD4+ T cell isolation kit (negative selection, STEMCELL) or the CD4+ T cell isolation kit (negative selection, Miltenyi Biotec). DNA was extracted from the isolated CD4+ T cells using the QIAamp DNA Mini or Micro kit according to the manufacturer’s instructions (Qiagen). At the time of plating with antigen, a baseline condition of ∼5M CD8-depleted PBMCs was set aside for CD4+ T cell isolation and DNA extraction as above, to assess baseline frequencies of clonotypes.

TCRβ repertoire sequencing of CD4+ T cell DNA from each well and data pre-processing were performed by the Johns Hopkins FEST and TCRΒ Immunogenomics Core Facility as previously described^114^. Processed data files were uploaded to the publicly available MANAFEST analysis web app (www.stat-apps.onc.jhmi.edu/FEST) to bioinformatically identify antigen-specific T cell clonotypes. The program uses only productive clones and for each clone, the program applies the Fisher exact test to compare the number of templates in a culture of interest and a reference culture. All clonotypes were subject to a 5-template lower threshold across all wells for consideration in the statistical analysis. The following analysis was performed separately to identify HIV-responding clonotypes and CMV-responding clonotypes. Clonotypes were deemed expanded if their abundance was significantly higher in the relevant well relative to a reference well using Fisher exact test with Benjamini-Hochberg correction for false discovery rate (FDR) p<0.05. Antigen-responsive clonotypes met the following criteria: (1) significantly expanded in the antigen-containing well (in at least two of three replicate wells) compared to the reference well (negative control or baseline) by Fisher’s exact test with Benjamini-Hochberg FDR, p<0.05, (2) was not significantly expanded in any of the negative wells performed in tandem compared to the reference culture by the same criteria, (3) significantly expanded in at least 2 of 3 antigen-containing wells compared to the reference well (negative control or baseline) with odds ratio >5, (4) if there was significant expansion in a negative well compared to reference, then the total abundance of the clonotype must be greater than the maximum of 9 or the baseline well template abundance in all three antigen-containing wells. Antigen-responsive clonotypes that met the following criteria were also included: (1) significantly expanded in at least one antigen-containing well compared to the reference well (negative control or baseline) by Fisher’s exact test with Benjamini-Hochberg FDR, p<0.05, (2) was not significantly expanded in any of the negative or positive wells performed in tandem compared to the reference culture by the same criteria, (3) significantly expanded in at least one antigen-containing well compared to the reference well (negative control or baseline) with odds ratio >5, (4) was not significantly expanded in any of the negative or positive wells performed in tandem compared to the reference culture with odds ratio > 5, and (5) its maximum frequency in an antigen-containing well divided by the sum of its maximum frequency in any other non-antigen well plus 0.001 to account for negative frequencies was >200.

## CMV IgG testing

The presence of CMV IgG was determined qualitatively using an indirect chemiluminescence immunoassay (LIAISON® CMV IgG, DiaSorin Inc) in the Johns Hopkins Medical Laboratories on cryopreserved serum samples from t0 or a sample prior to t0 in all longitudinally sampled participants and from the cross-sectionally sampled PWH on ART. The CMV serostatus of the cross-sectionally sampled HIV-negative people was previously published^53^.

## Downsampling rank abundances and computing ecological metrics

To produce rank-abundance distributions (curves), we arrange all clonotypes by their rank (*r* ∈ [1, *R*]) from largest to smallest abundance *a*(*r*) where *R* is the richness or total number of unique species in the sample. We then assume that each clones’ proportional abundance is given by its observed abundance *p*(*r*) = *a*(*r*)/*N* where *N* is the original experimental sample size (**Figure S3**). Note this means that many sequences are observed with the same abundance, such that their rank must be randomly assigned. Using a multinomial distribution, we then find the downsampled abundance of each clone as *M*(*p*(*r*), *s*) where *s* is the resampled size.

After downsampling to the same sizes, we chose to compare distributions summarized with Hill numbers, ecological metrics that can be derived from a mathematically unified equation (**Supplementary Methods 1**) where the *x*-th Hill number is 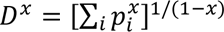 given the clone proportional abundances *p*_*i*_ where *i* indicates individual clonotypes. Further, we normalized these metrics to *D*^*x*^/*N*\* where *N*\* is the common re-sample size such that all metrics ranged from zero to one.

### Fitting to rank abundances

In the main text, we present the results of fitting with two models: the single and double power law models. Other models were tested including negative binomial, log-series, and exponential power law (see **Supplementary Methods 1**) but none worked as well as these two options and were deemed unnecessarily complex. To fit these models, we sample a model to the same *N* as the experimental data and estimated the model parameters that minimized (using *scipy.optimize* package) the root-mean-square error between the cumulative proportional abundances of the data and the model. We also used a version of model fitting that minimized the Kolmogorov-Smirnov statistic and found similar results.

### Simulating reservoir population decline from TCRβ clone data

We began by assuming that HIV DNA is at the population average level across the IPDA data, *H*\*(0)=800 intact and *H*(0)=3000 defective proviruses per million CD4+ T cells, respectively (**Figure 5F**). Then, beginning at t0 for each participant data set we randomly choose CD4+ T cell clones, add their abundance, and continue until reaching *H*(0) cells. The number of rmCD4+ T cells in a sample is the sum of all clone abundances *T*(*t*) = ∑_*i*_ *T*_*i*_(*t*) and then the number of HIV proviruses (inclusive of intact, but likely mostly defective) is accordingly *H*(*t*) = ∑_*j*_ *T*_*j*_(*t*) where *j* are the indices of TCRβ clones that are infected --note for now we assume that all members of a selected TCRβ clone harbor proviruses. Thus, we calculate the proportion of total HIV DNA per million CD4+ T cells at the first time point *f*_0_ = *H*(*t*_0_)/*T*(*t*_0_) × 10^6^.

Then, we examine the abundance of each of the *j* clones carrying HIV DNA at following time points. Importantly, we use TCRβ sequencing inclusive of naïve cells to properly estimate the denominator *T*(*t*_1,2_) size, as fractions of naïve CD4+ T cells can change in PWH on ART. At each of the following time points, we calculate the proportion of HIV DNA from clones that were observed in t0 related to the total CD4+ T cells from all clones (*f*_1_, *f*_2_). This procedure numerically approximates how the proportion of HIV DNA per million CD4+ T cells would change over time if it was purely governed by TCRβ clonal dynamics. Finally, from these proportions, we estimated the rate of change as in the modeling of IPDA data, assuming an exponential decay model and fitting using log-linear mixed effects. We replicated this procedure 10 times for each of the 4 Ctl participants who had naïve and memory TCRβ data available. Then we calculated a mean rate of change for each participant and then the population mean and 95% CI, which could then be compared to our IPDA estimates.

## Simultaneous CD4 carrier and HIV passenger mechanistic model

A complete mathematical description of the mechanistic model is provided in the **Supplementary Methods 1**. We use a tau-leap algorithm^115^ to stochastically simulate a set of ordinary differential equations (time step Δ*T*=30 days, simulations over 10 years) where each TCRβ clone has an equation with a randomly selected but balanced birth and death rate. Initial clone abundances were drawn from a distribution, which was varied (constant, uniform, exponential, power-law), as were assumptions about the distribution of balanced proliferation and death rates (constant, uniform, exponential). We compared model output to comprehensive experimental data from **Fig 1-4** in this manuscript, determining the optimal model that could match all data by accepting models that fell within experimental ranges for CD4+ T cell and HIV DNA population sizes and longitudinal dynamics and clonality of intact and defective HIV DNA and CD4+ TCRβ sequences as well correlations between TCRβ clone sizes and fold-changes and the distribution of TCRβ clone expansions/contractions. The metrics that each of 7 models succeeded and failed to match are documented in the **Supplementary Methods 2** file.

## Supplementary Tables and Figures

**Supplementary Table 1. Detailed participant and sample characteristics. [Separate file]**

Abbreviations: AA, African American; ABC, abacavir; ART, antiretroviral; CMV, cytomegalovirus; DTG, dolutegravir; EFV, efavirenz; EVG/c, elvitegravir/cobicistat; FTC, emtricitabine; JHHCC, Johns Hopkins HIV Clinical Cohort; m, months; LPV/r, lopinavir/ritonavir; ND, not done; NFL, near-full length; PWH, people living with HIV; RPV, rilpivirine; t0, t1, etc, study timepoint 1, 2; TCRβ, T cell receptor beta chain; TDF, tenofovir disoproxil fumarate; Unk, unknown; W, White; ZDV, zidovudine; 3TC, lamivudine.

**Supplementary Figure 1.**
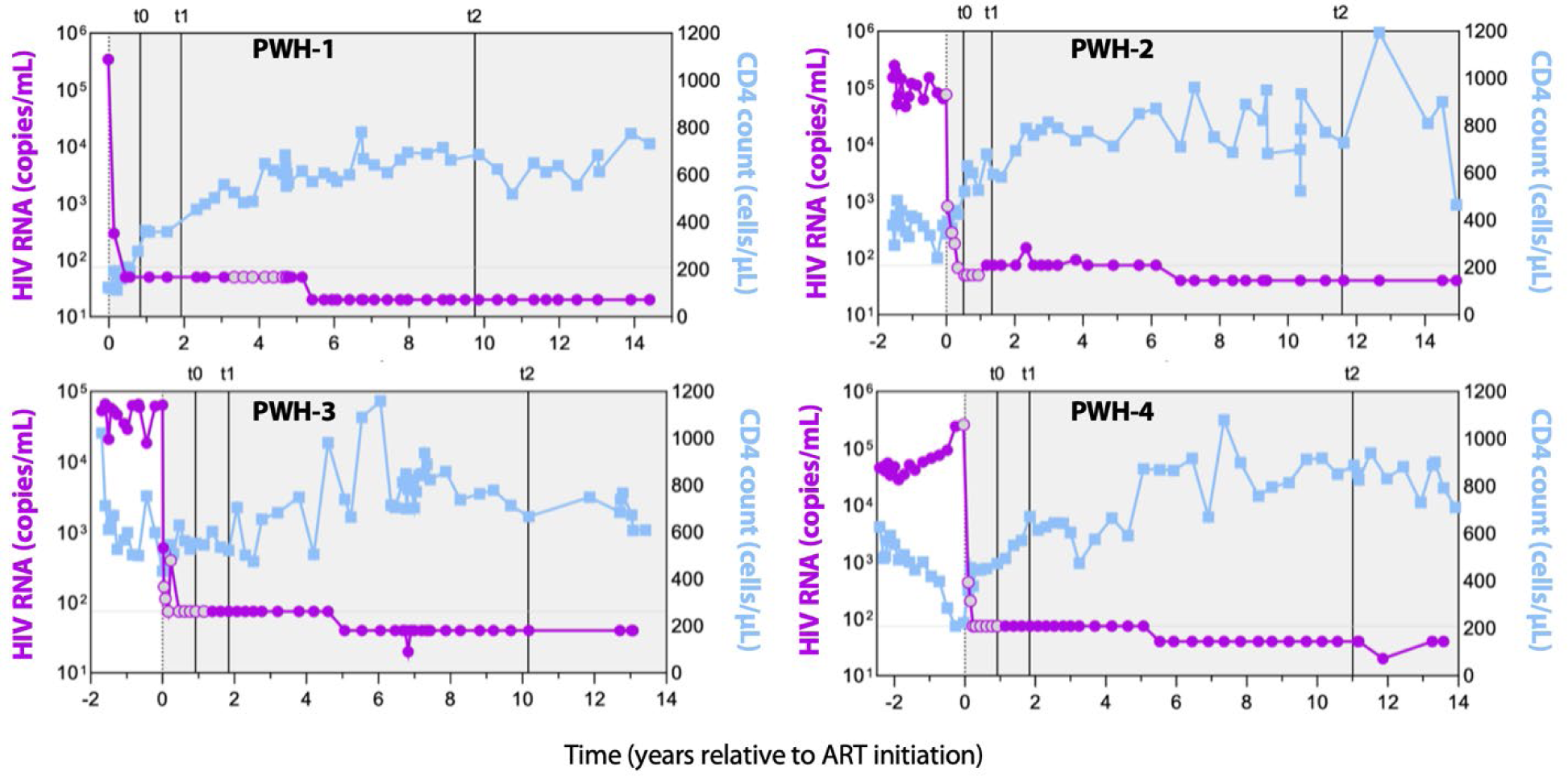
Longitudinal participant viral loads and absolute CD4+ T cell counts. HIV-1 RNA copies/mL (purple) and absolute CD4+ T cells/µL (blue) in the peripheral circulation of the four PWH on ART sampled longitudinally in this study. Grey shading indicates time on ART. Sample time points t0, t1, and t2 are shown for each participant. Resting memory CD4+ TCRβ repertoires were sequenced at t0, t1, and t2. Near-full length HIV proviruses isolated from resting CD4+ T cells were sequenced at t1 and t2.

**Supplementary Figure 2.**
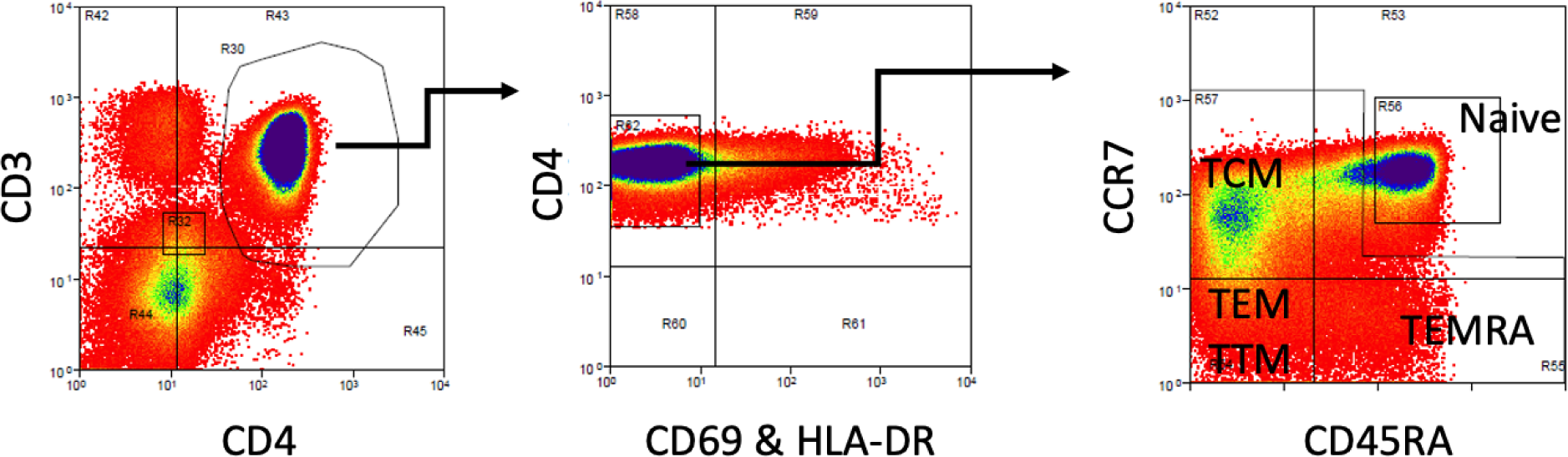
Gating strategy for FACS sorting of resting memory CD4+ T cells. A dump channel included CD8+, CD14+, CD16+, CD20+, and non-viable cells (Methods). Selected cells were CD8-CD14-CD16-CD20-CD3+CD4+CD69-HLA-DR- and either CCR7- or CCR7+CD45RA-(collected together). TCM, T central memory cells, TEM, T effector memory cells, TTM, T transitional memory cells, TEMRA, T effector memory cells re-expressing CD45RA.

**Supplementary Figure 3.**
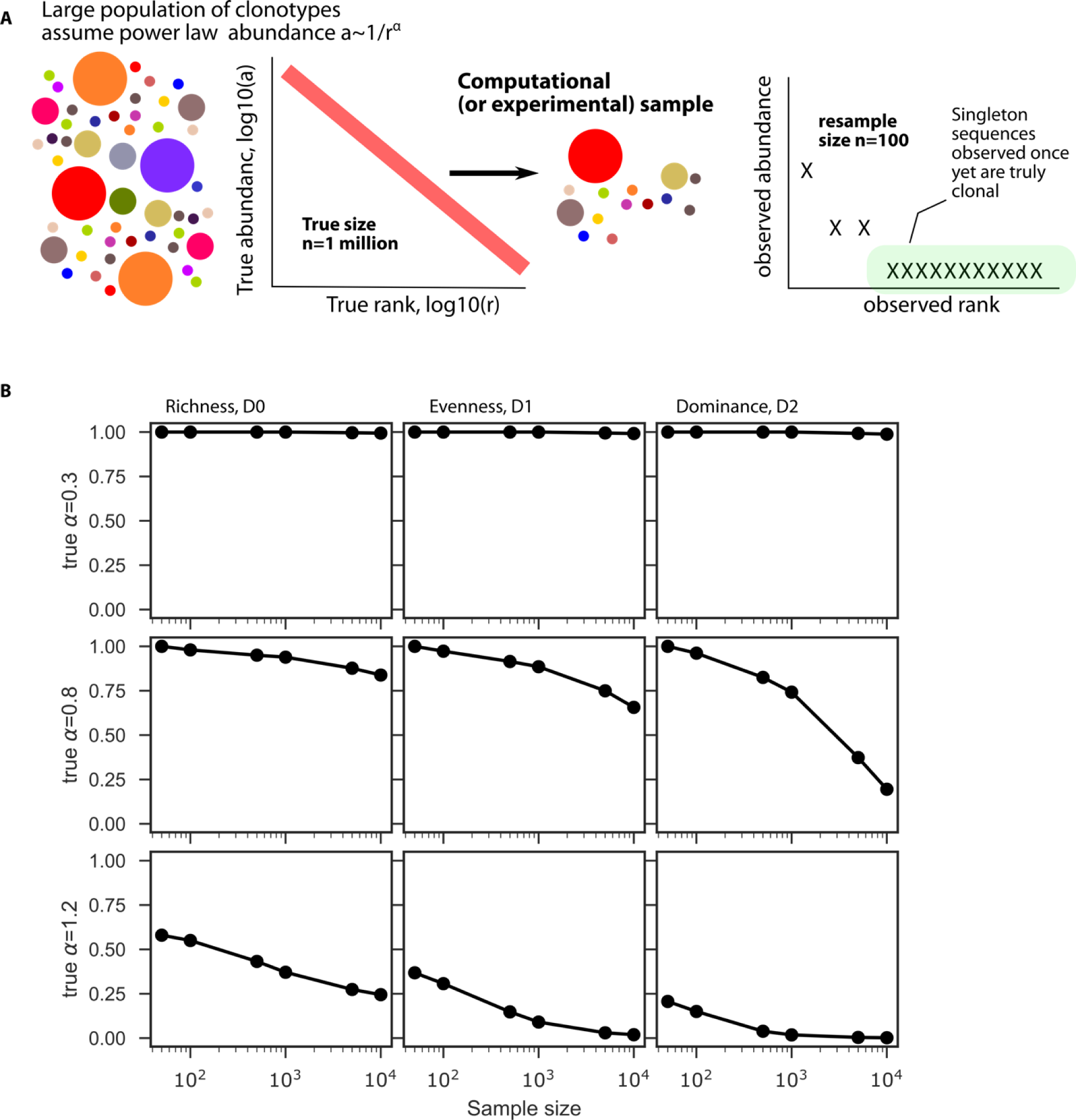
Simulation study to illustrate influence of sample size on ecological metrics. A) Model schematic, we assume clones follow a strict power law rank abundance relationship with exponent alpha, *a*(*r*)∼*r*^−*α*^ and true size (number of cells) n=10^6^ and then resample these distributions to a lower sample size. Resampled rank abundance curves (here n=100) can contain many observed singletons that are truly clonal in the original distribution. B) Three “Hill” diversity metrics computed from different underlying power law exponents (rows) at varying sample sizes (x-axis) illustrate that observed clonality depends on sample sizes, and this dependence grows stronger with alpha values.

**Supplementary Figure 4.**
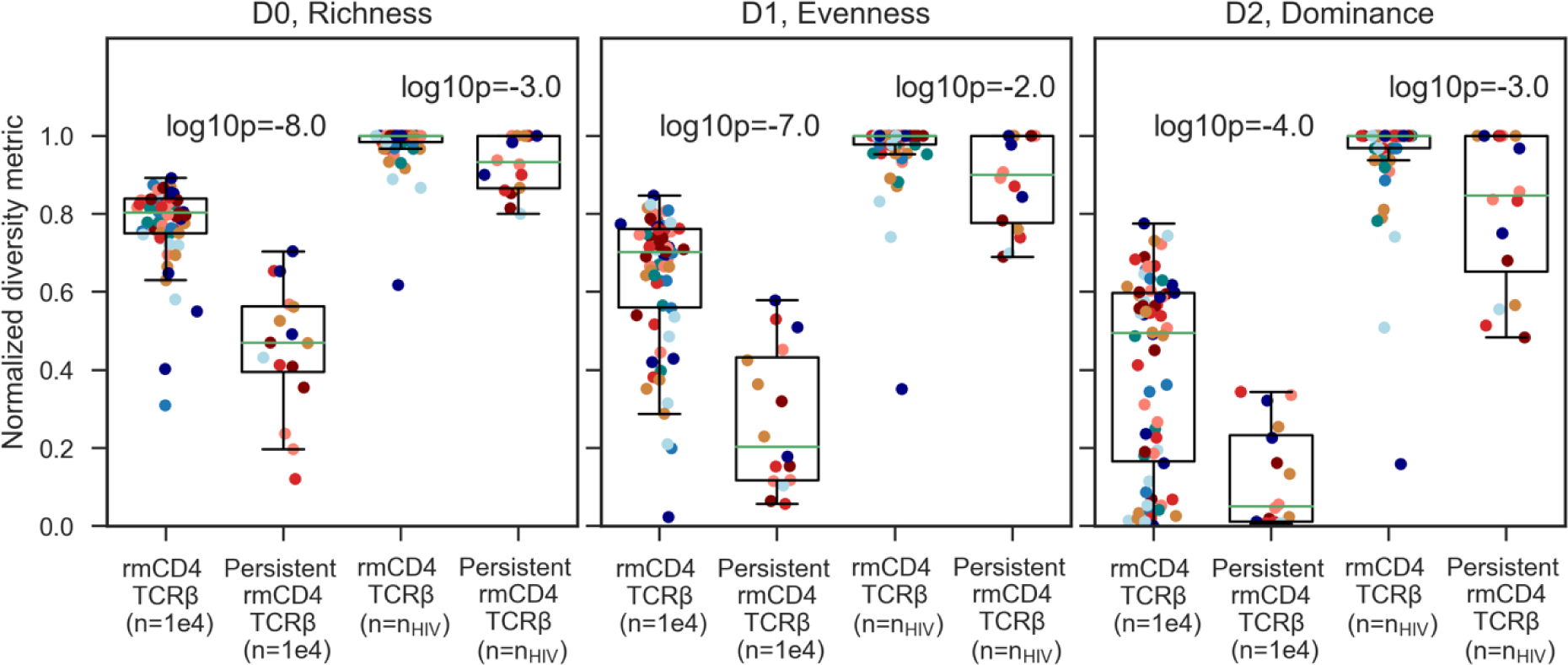
Comparison of normalized diversity metrics for TCRβ and persistent TCRβ at resampled sizes for typical TCRβ and HIV proviruses. Ecological diversity metrics of rmCD4 TCRβs and persistent rmCD4 TCRβs downsampled to *n*=10^4^ (left) and downsampled to the sample size of HIV proviruses from that participant time point (right). HIV proviruses were not sequenced at t0, so we set the downsampled size for TCRβ sequences at t0 to *n*_*HIV*_ =60. P-values indicate paired differences using a one-sided Mann-Whitney test.

**Supplementary Figure 5.**
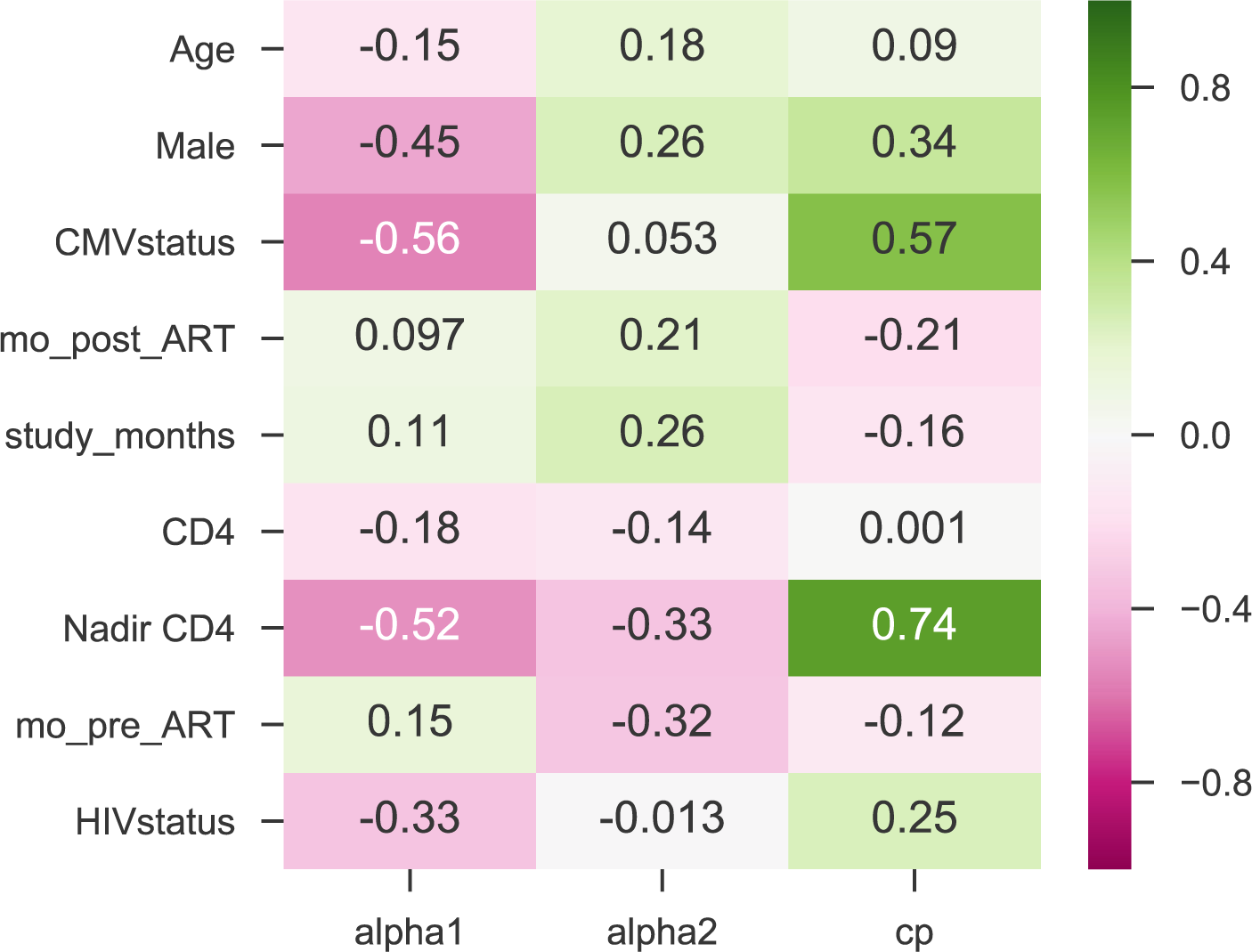
Spearman correlation coefficients computed between power law slope parameters and clinical variables. We assessed associations between large clone slope (alpha1), smaller clone slope (alpha2), and change point of that slope (cp, the rank where the slope changes) and 9 clinical variables (y-axis). Associations were deemed significant with a Bonferonni correction threshold of p<0.05/27=0.002, which occurs in this data set when correlation coefficients have absolute values above 0.4. Thus, significant associations are observed between alpha1 and Male sex, CMV status, and Nadir CD4 count; and between cp and CMV status and Nadir CD4 count.

**Supplementary Figure 6.**
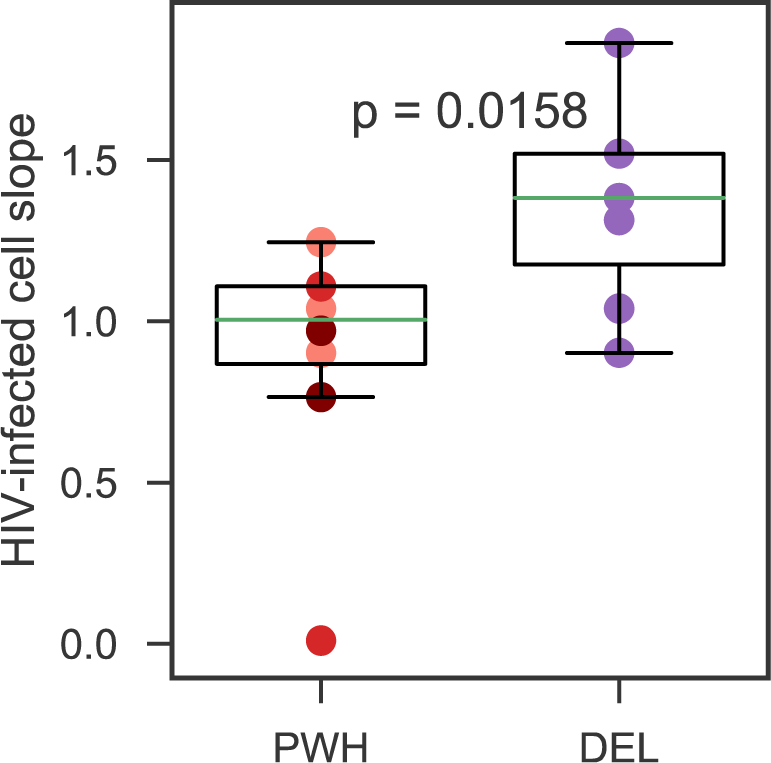
Power law slope for HIV-infected cells from two cohorts. DEL cohort HIV reservoir cells (*Env* sequences from intact, inducible proviruses) had significantly higher slopes compared to the PWH cohort HIV-infected cells (near-full length sequences from intact and defective proviruses) (p=0.02, 1 sided Mann-Whitney U test) indicating higher clonality.

**Supplementary Figure 7.**
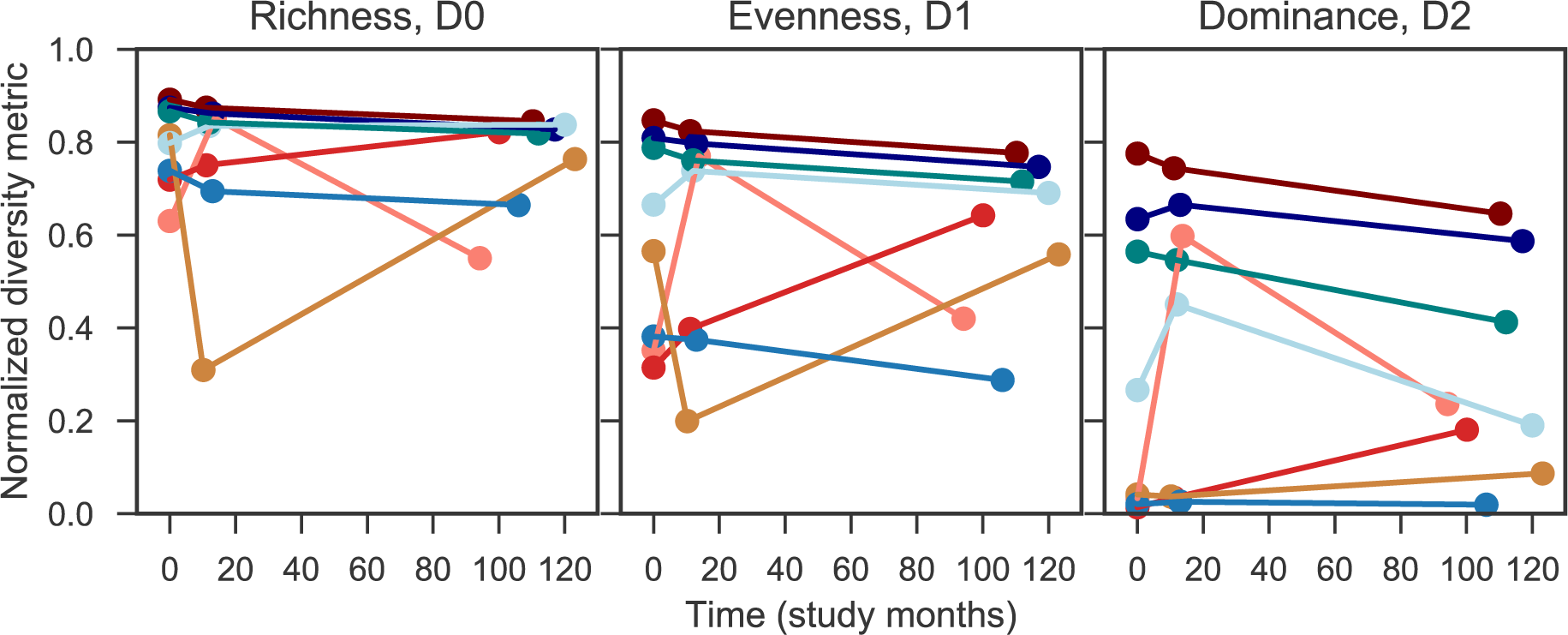
Longitudinal ecological metrics for TCRβ data from PWH and Ctl groups. No significant trends are detected via linear mixed effects model (p>0.05 on slope). Study months indicates time relative to t0. Individuals denoted by different colors, PWH in reds and oranges and HIV-seronegative people in blues and greens.

**Supplementary Figure 8.**
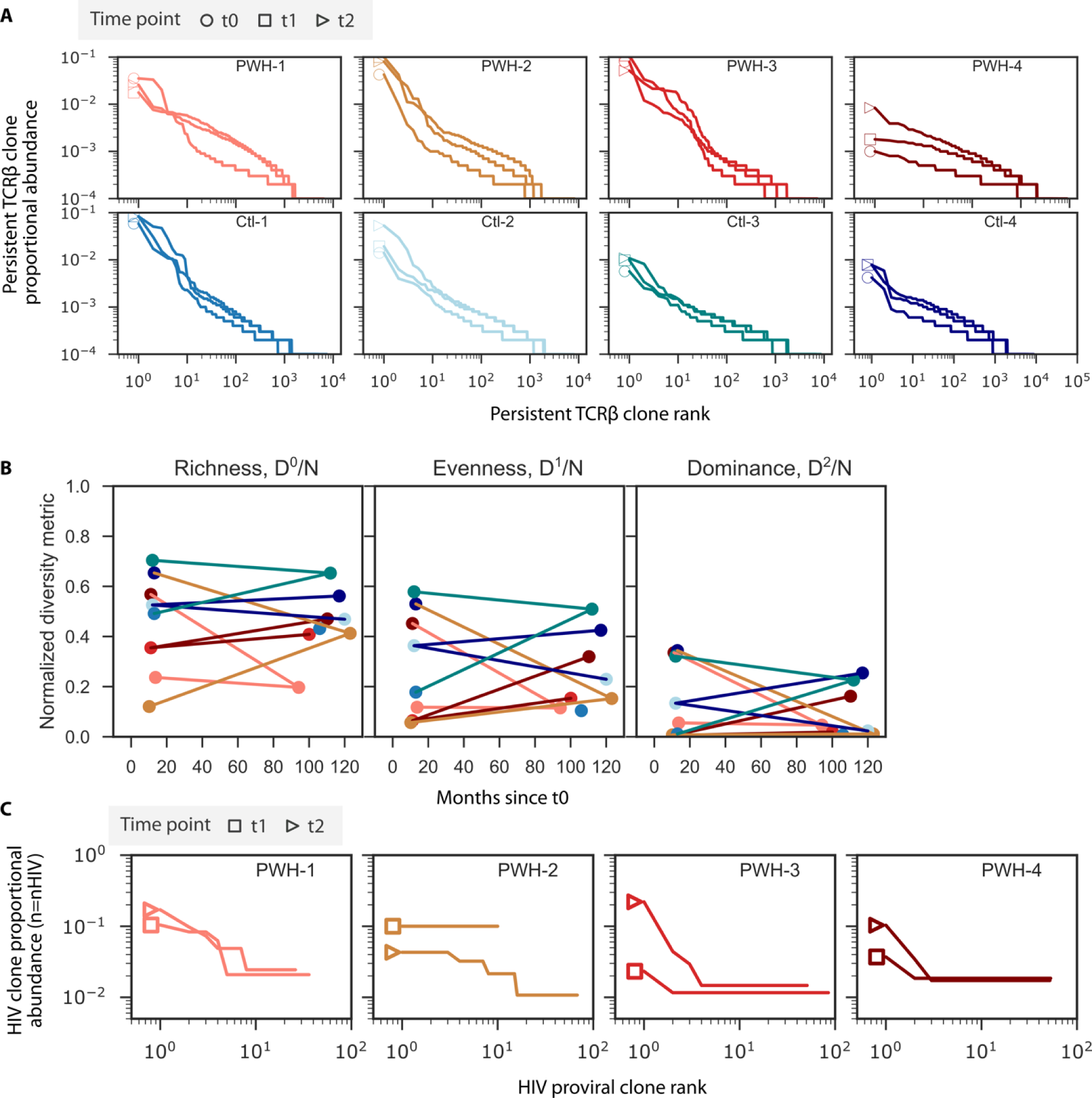
Longitudinal persistent TCRβ compared with longitudinal HIV provirus rank abundance and ecology. A) Proportional rank abundance distributions of persistent TCRβ at all three time points. B) Diversity metrics from longitudinal PWH (reds) and Ctl cohorts (blues). Pearson correlation coefficients indicate no trends over time (p>0.05). C) Proviral proportional rank abundance distributions of raw proviral data (not resampled) at t1 and t2 illustrate visually how clonality increases in 3/4 participants (compare triangles and squares) and in PWH 3 the change is unclear because t1 had only n=10 samples, all singletons.

**Supplementary Figure 9.**
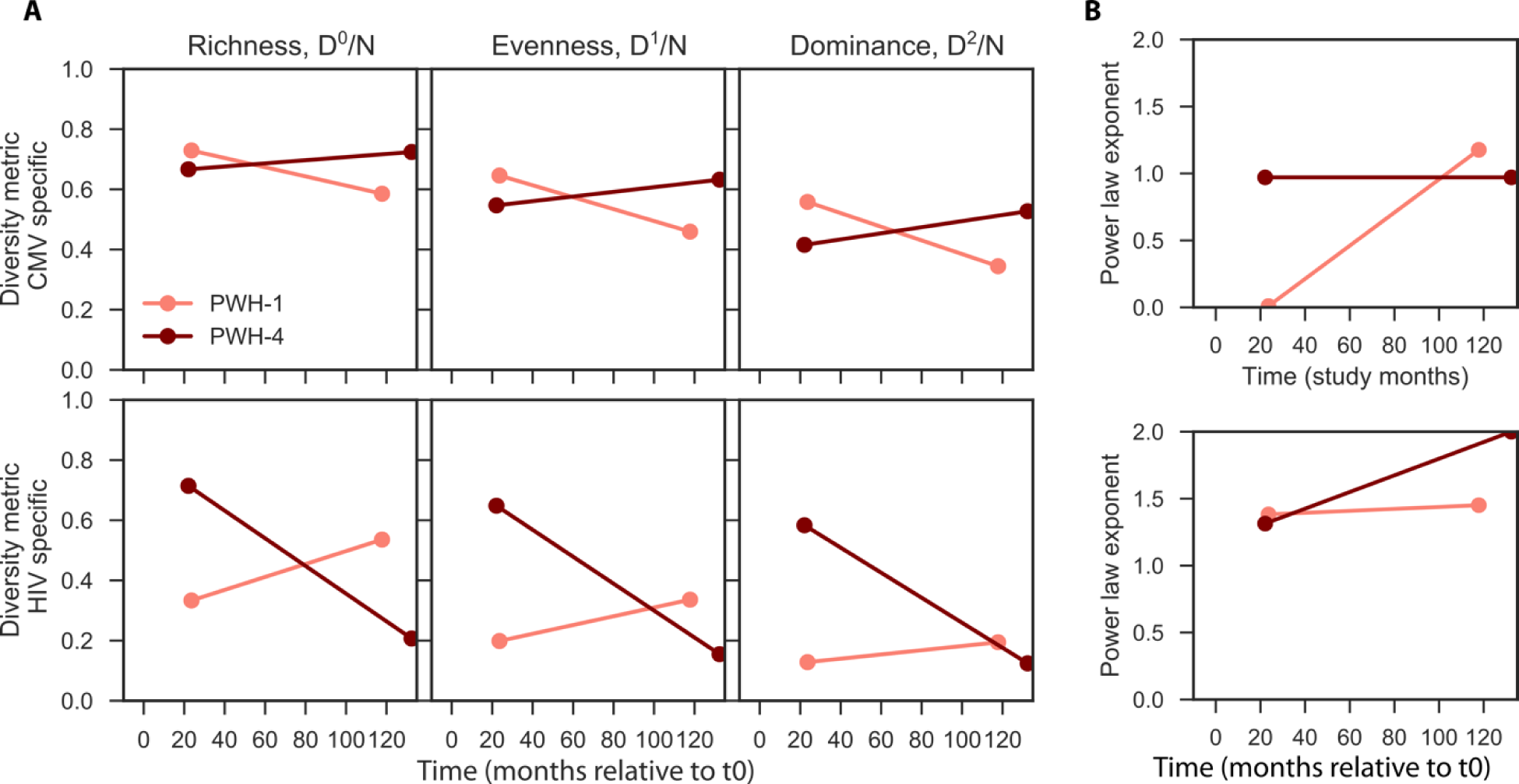
Longitudinal ecological metrics (A) and power law exponents (B) estimated from antigen specific TCRβ data. Across 2 participants (PWH-1 and PWH-4, colors correspond across all panels) no significant trends are detected via pearson correlation coefficient (p>0.05 on slope). Study months indicates time relative to t0.

**Supplementary Figure 10.**
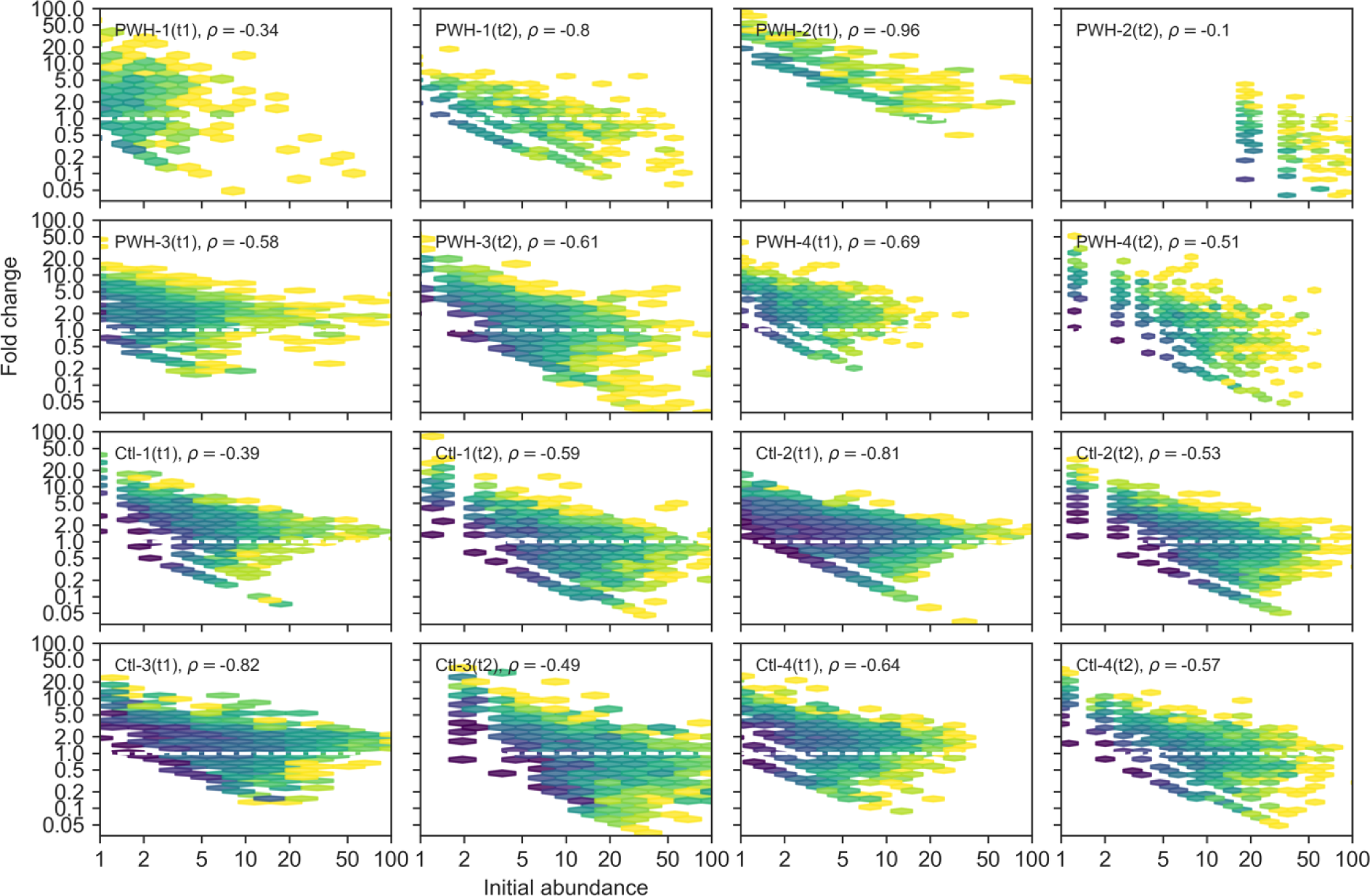
Correlation between initial mCD4 TCRβ clone size and fold-change in clone size across for all PWH and Ctl. For each participant time point (t1 and t2), the initial abundance was determined at the prior time point (t0 and t1, respectively). Then the fold change of clonotypes found at both time points is shown. All data are derived from resamples where each time point has the same n=1e4. Darker regions indicate higher density of points. Spearman correlation coefficients are annotated on each panel, indicating significant negative trends such that larger initial sizes predict contraction and smaller initial sizes predict expansion.

## Notes

### Author Declarations

Ethics committees/IRBs of the Johns Hopkins University School of Medicine, the University of California San Francisco, and the University of Pennsylvania gave ethical approval for this work.

