## Supplementary material for "Mild HIV-specific selective forces overlaying natural CD4+ T cell dynamics explain the clonality and decay dynamics of HIV reservoir cells": Supp Methods 1

### Contents

**1 Classical ecological metrics** **1**

**2 Modeling and analysis of T cell receptor (TCR$\beta$) rank abundance distributions** **3**

2.1 Mathematical models for rank abundance distributions . . . . . . . . . . . . . . . . 4

2.2 Approximating the normalizing constant of rank abundance distributions by integration 5

**3 Mechanistic model of simultaneous HIV and TCR$\beta$ clonal dynamics.** **6**

### 1 Classical ecological metrics

To summarize distributions of abundances of genetic sequences it is useful to imagine each unique sequence as its own species and invoke work from animal ecology. One of the most basic ecological metrics is the richness $R$, the number of unique species. Because our sampling is necessarily incomplete, we sometimes specify the *observed* richness $R^{obs}$. Sample size is denoted $N$. We refer to the distribution of observed species abundances $a_s$, denoting every species $s$ by its relative abundance $p_s = a_s/N$

We then often visualize species abundances as a *rank abundance* curve: each species is given a rank $s \to r$, ranked from largest to smallest in terms of sampled abundance $a_r$. Ranks are discrete, $r \in [1, 2, \ldots]$, and the maximum rank is the observed richness $R^{obs}$. When species have the same abundance, they are still given unique ranks but randomly ordered. That is, we can have $a_{r+1} = a_r$.

A natural method to summarize the relative species abundance distribution uses the Shannon entropy $\mathcal{S} = -\sum_r p_r \ln p_r$, which can be normalized to the theoretical maximal entropy (uniform distribution) given the sample size. That is, if all $p_r = 1/R$, then $\mathcal{S}_{max} = -\sum_r (1/R)\ln(1/R) = \ln R$. The ecological evenness (now scaled from 0-1 from most to least uneven) is

$$E = \mathcal{S}/\ln R \tag{S1}$$

A further general way to quantify diversity from relative abundances via Hill[1] is through inverse $p$-norms, or inverse weighted power means. For all values $x_i$ in a set, the weighted power mean is: $M_P(x_i; w_i) = (\sum_i w_i x_i^P)^{1/P}$, where $w_i$ is the weight.

[1] Hill 'Diversity and evenness: a unifying notation and its consequences'. Ecology. 54 (2): 427–432 (1973)

To define generalized diversity measures, we take the inverse of the power mean, call $P = q - 1$, and set $w_i = p_r$. Thus ${}^qD := \left(\sum_r p_r p_r^{q-1}\right)^{-1/(q-1)}$ and finally,

$$ {}^qD = \left(\sum_r p_r^q\right)^{1/(1-q)} \tag{S2} $$

Following through the first 3 Hill numbers, $q = 0$ would admit the weighted harmonic mean for the power mean, and for our definition results in the richness.

$$ {}^0D = \sum_r 1 = R \tag{S3} $$

The second Hill number $q = 1$ would is the weighted geometric mean, is not derived trivially, one can rewrite in terms of exponentials $\exp\ln({}^1D)$ and apply l'hôpital's rule $\lim_{x\to c} f(x)/g(x) = \lim_{x\to c} \partial_x f(x)/\partial_x g(x)$ to calculate the expression in the limit $q \to 1$, which interestingly results in the exponential of the Shannon entropy

$$ {}^1D = \exp(\mathcal{S}) \tag{S4} $$

The third Hill number, $q = 2$, which would represent the weighted arithmetic mean, simplifies more easily

$$ {}^2D = \left(\sum_r p_r^2\right)^{-1} = \frac{1}{\lambda} \tag{S5} $$

where $\lambda$ is known as the Simpson index in ecology and the Herfindahl-Hirschman index (HHI) in economics. It can be roughly interpreted as the total probability of sampling and finding the same clone twice from sampling two cells (i.e. the probability of sampling rank $j$ twice is the product $p_j^2$) summed over all ranks.[2] Naturally, the probability that any two sampled cells are different is $1 - \lambda$, which is the Gini-Simpson index.

Finally, if $q \to \infty$, we can rewrite as

$$ {}^qD = \left(p_1^q \sum_{r=2} \left(\frac{p_r}{p_1}\right)^q\right)^{1/(1-q)} \tag{S6} $$

such that all entries in the sum go to zero after exponentiation, and we are left with $p_1^{q/(1-q)}$, which reduces to $1/p_1$. Thus

[2] Caution, this interpretation assumes replacement or that sampling one cell does not meaningfully reduce the relative abundance, an approximation that is fine for large data sets

$$^{\infty}D \sim \frac{1}{p_1} \tag{S7}$$

the inverse relative abundance of the most abundant clone.[3] Through this, we see that as the Hill numbers rise, the intuition is that the diversity reflects more and more the larger clones.

Overlap (per Morisita[4]), can be defined as the total probability of getting the same sequence twice in 2 samples divided by the Simpson's index within each sample (i.e. the probability of getting the same sequence twice in each sample)

$$I_\sigma = \frac{2\sum_s p_s(1)p_s(2)}{\sum_s p_s(1)^2 + \sum_s p_s(2)^2} \tag{S8}$$

#### 2 Modeling and analysis of T cell receptor (TCR$\beta$) rank abundance distributions

We tested several mathematical models against T cell receptor sequence distribution data. To fit mathematical models, we perform several steps.

First calculate the proportional rank abundance

$$p(r) = \frac{a^{obs}(r)}{\sum_r a^{obs}(r)} \tag{S9}$$

Resample to the same size $N$ for all individuals and time points, such that the probability of drawing each sequence is assumed to be reflected by the original experimental sample.

$$a_N(r) = \mathcal{M}(r; N, p(r)) \tag{S10}$$

We then tested several mathematical models $m$ against these resampled rank-abundances to see which one optimally captured these data. We define optimal as the minimization of the root-mean-squared ($RMS$) error between the model and data distributions when cast as cumulative proportional rank abundances (or cpa, denoted as $c_m$ and $c_d$ for the model and data, respectively).

$$RMS = \left\{ \sum_{r=1}^{R^{max}} \left[c_m(r) - c_d(r)\right]^2 \right\}^{1/2}. \tag{S11}$$

Note this calculation was adjusted to only score fit against non-singletons by choosing an upper limit $R^{max}$ because for singletons the assumption that the sampled abundance is proportional to the true abundance is very weak. Also note that this procedure amounts to a similar approach to minimizing

[3]Note if there are multiple clones $n_1$ of the same abundance (the maximum abundance) this reduces to $\frac{1}{n_1 p_1}$.

[4]Morisita, M. I$_\sigma$-Index, a measure of dispersion of individuals. Res Popul Ecol 4, 1-7 (1962).

the Kolmogorov-Smirnov statistic (the largest deviation between the cpas). However, because of the logarithmic nature of the ranks, we felt the distribution was more representative if the error was averaged across the distribution, rather than penalizing the worst part.

Computationally, we found the minimum using Python and the `scipy optimize` package.

#### 2.1 Mathematical models for rank abundance distributions

We began with a single power law because we used this model for HIV sequence rank abundances previously[5].

$$m(r) = \psi(\alpha, R) r^{-\alpha} \tag{S12}$$

where we have

$$\psi(\alpha, R) = \sum_{s=1}^{R} s^{-\alpha}, \tag{S13}$$

is a normalizing constant that depends on the model parameter $\alpha$, which we refer to throughout as the power law exponent, and crucially also depends on the assumed richness of the model $R$.

The data were not as simply log-log linear as prior data, so we also tried a 'double' or 'two-phase' power law model

$$m(r) = \psi(\alpha_1, \alpha_2, R) \left[ r^{-\alpha_1} + \phi r^{-\alpha_2} \right] \tag{S14}$$

which again requires an analogous normalization factor.

Determining the change point $r_{cp}$, or the rank when the second phase becomes the bigger contributor to the rank abundance can be accomplished by setting:

$$r_{cp}^{-\alpha_1} = \phi r_{cp}^{-\alpha_2} \tag{S15}$$

which can be simplified via logs $(\alpha_2 - \alpha_1) \ln r_{cp} = \ln \phi$ leading to

$$r_{cp} = \phi^{\frac{1}{\alpha_2 - \alpha_1}} \tag{S16}$$

Following McGill et al. Ecol Lett 2012 we also tried a few interesting distributions: log-series, derived from the Taylor-series expansion of the logarithm

[5] Reeves et al. A majority of HIV persistence during antiretroviral therapy is due to infected cell proliferation. Nature communications 9 (1), 4811 (2018)

$$m(r) = \psi(k, R) \frac{1}{\log(1-k)} \frac{k^r}{r} \tag{S17}$$

Negative binomial:

$$m(r) = \binom{r+n-1}{n-1} p^r (1-p)^k \tag{S18}$$

What we call an exponential power-law:

$$m(r) = \psi(A, R) \exp r^{-A} \tag{S19}$$

#### 2.2 Approximating the normalizing constant of rank abundance distributions by integration

We observe that certain model parameter combinations are impossible. For example, for a given power law exponent, the richness is constrained below a certain value for a given reservoir size. This observation has been considered previously in ecology under the terminology of 'feasible sets' (Locey et al 2013). Because the partial sum $\psi(\alpha, R)$ has no analytical solution we are aware of, we approximate the sum using an integral for large $R$. That is, we use

$$\psi(\alpha, R) = \sum_{s=1}^{R} s^{-\alpha} \approx \int_1^R ds\, s^{-\alpha} = \frac{R^{1-\alpha} - 1}{1-\alpha} \tag{S20}$$

to compute an approximate rank-abundance distribution

$$\tilde{a}(r) = \left| \frac{L\, r^{-\alpha}}{\psi(\alpha, R)} \right| \tag{S21}$$

Then, we solve for the maximum richness by considering the rounding process and finding the maximum first value of $r$ for which $a(r) = 0$, which is when $L\, r_{max}^{-\alpha}/\psi(\alpha, R) \leq 1/2$. We use the condition of equality, to solve for $r_{max}$, which results in

$$r_{max} \approx \exp\left[ -\frac{1}{\alpha} \ln\left( \frac{\psi(\alpha, R)}{2L} \right) \right]. \tag{S22}$$

Note that the expression still depends on the initial $R$ of the model. We checked that this approximation was reasonable in our parameter ranges by directly computing the maximum richness using the rank-abundance without approximation. This comparison is shown in **Supplementary Fig 2**. Some combinations do lead to $r_{max} > L$, in which case we set $r_{max} = L$.

### 3 Mechanistic model of simultaneous HIV and TCR$\beta$ clonal dynamics.

We built a multi-clonotype model of the TCR repertoire which includes a subset of HIV infected cells. Our general model is a system of differential equations with the form

$$\dot{T}_i = (b_i - d_i)T_i + \psi \tag{S23}$$

where we allow TCR's can emerge from new naive T cells which is modeled with the rate $\psi$ but found we could generally set this rate $\psi = 0$.

The system of equations is solved stochastically using the $\tau$-leap approach [Gillespie et al. 2001]. For example, at each time step $\Delta t$, we solve

$$\Delta T_i = \wp(b_i T_i \Delta t) - \wp(d_i T_i \Delta t) \tag{S24}$$

where $\wp$ is Poisson-distributed for each of the clones.

We made different assumptions about the initial conditions of clone sizes, constant, uniform, exponential, and power-law distributed.

We made different assumptions about the distribution of birth ($b_i$) and death ($d_i$) rates of the $i$-th clone by drawing from a distribution $b_i \sim p(b)$ which could be constant, uniform, or exponential, i.e., $p\left(b\right) = \lambda^{-1} e^{-\lambda b}$ with average rate $\lambda \sim 0.7$ from experimental data (Fig 5).

Then, to enforce global homeostasis, we force the deterministic derivative of each clone size to be zero ($\dot{T}_i = 0$), or that $d_i = b_i$.

Next we incorporate HIV DNA into randomly chosen TCR clones and assigning a fraction of the clone cells to harbor HIV DNA:

$$H_j\left(0\right) = f \times T_i(0) \tag{S25}$$

with the assignment $j \to i$. We use our own estimates of the frequency of HIV integration (Fig 5 in the main manuscript) to set 8000 total and 300 intact copies of HIV DNA per million CD4. We use a further subset $k$ that have intact HIV DNA.

Then, the dynamics of the $j$-th HIV infected cell follow

$$\dot{H}_j = (b_j - d_j - \xi_j) f T_j \tag{S26}$$

where the key difference is the impossibility of creating new cells due to thymic production (i.e. no $\psi$ term), but also the additional death term $\xi_j$ that models a negative selection against HIV clones. We ultimately found $\xi_k^*$ was required to model intact proviruses.
