## Supplementary material for "Mild HIV-specific selective forces overlaying natural CD4+ T cell dynamics explain the clonality and decay dynamics of HIV reservoir cells": Supp Methods 2

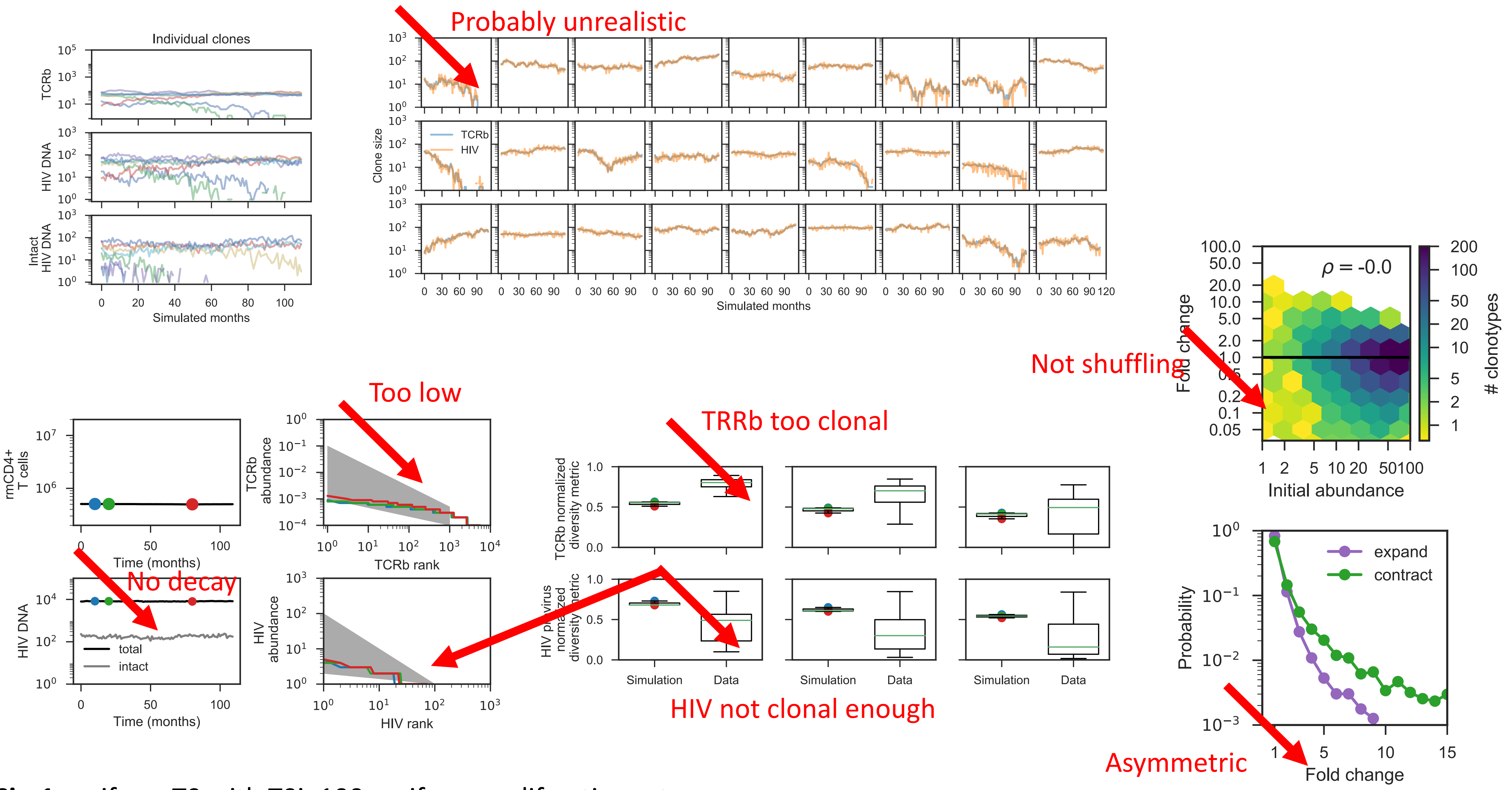


**Sim1**: uniform T0 with T0i=100, uniform proliferation rates, no reemergence, never redraw rates, no selection on proviruses, full coverage of TCR by proviruses

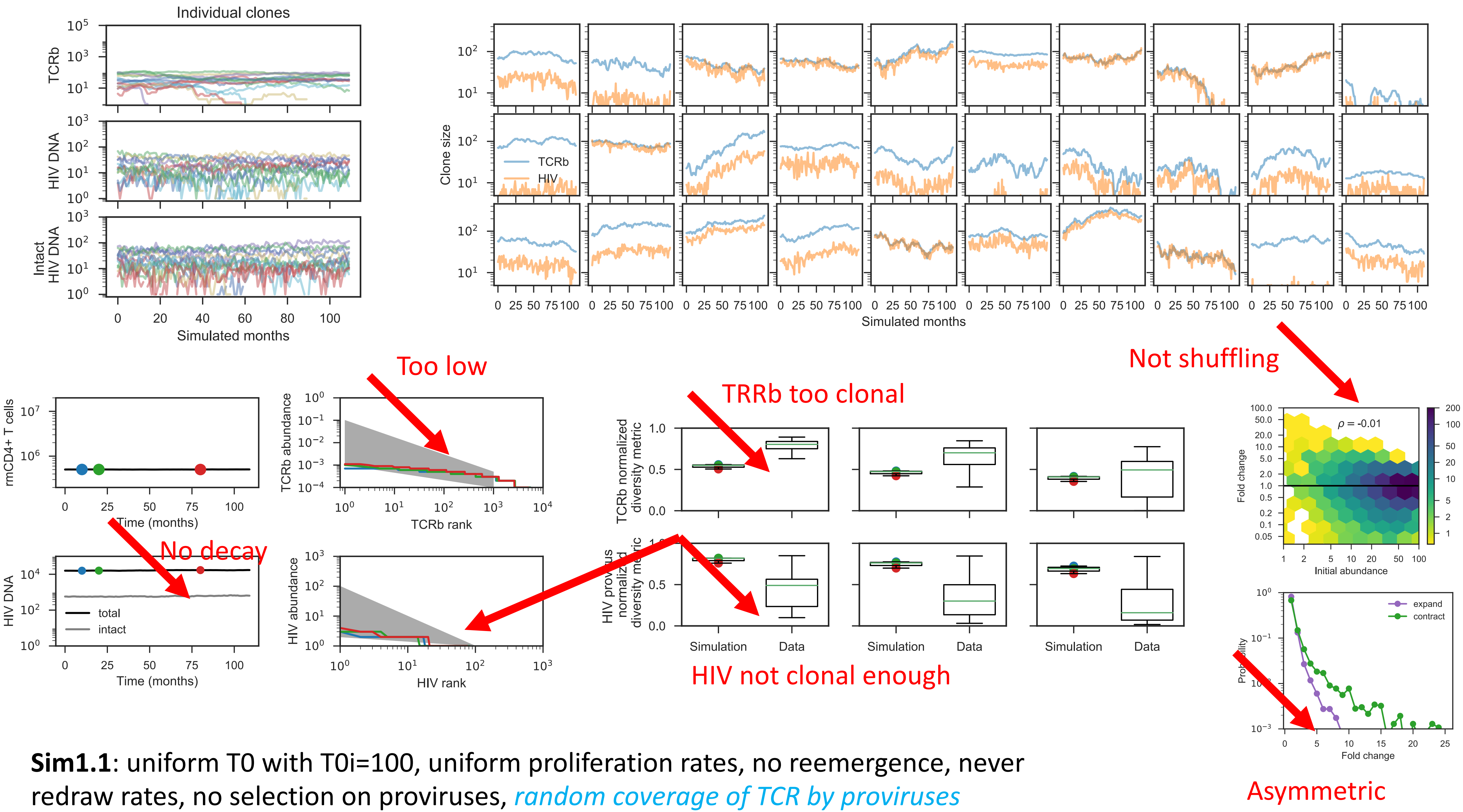


**Sim1.1**: uniform T0 with T0i=100, uniform proliferation rates, no reemergence, never redraw rates, no selection on proviruses, *random coverage of TCR by proviruses*

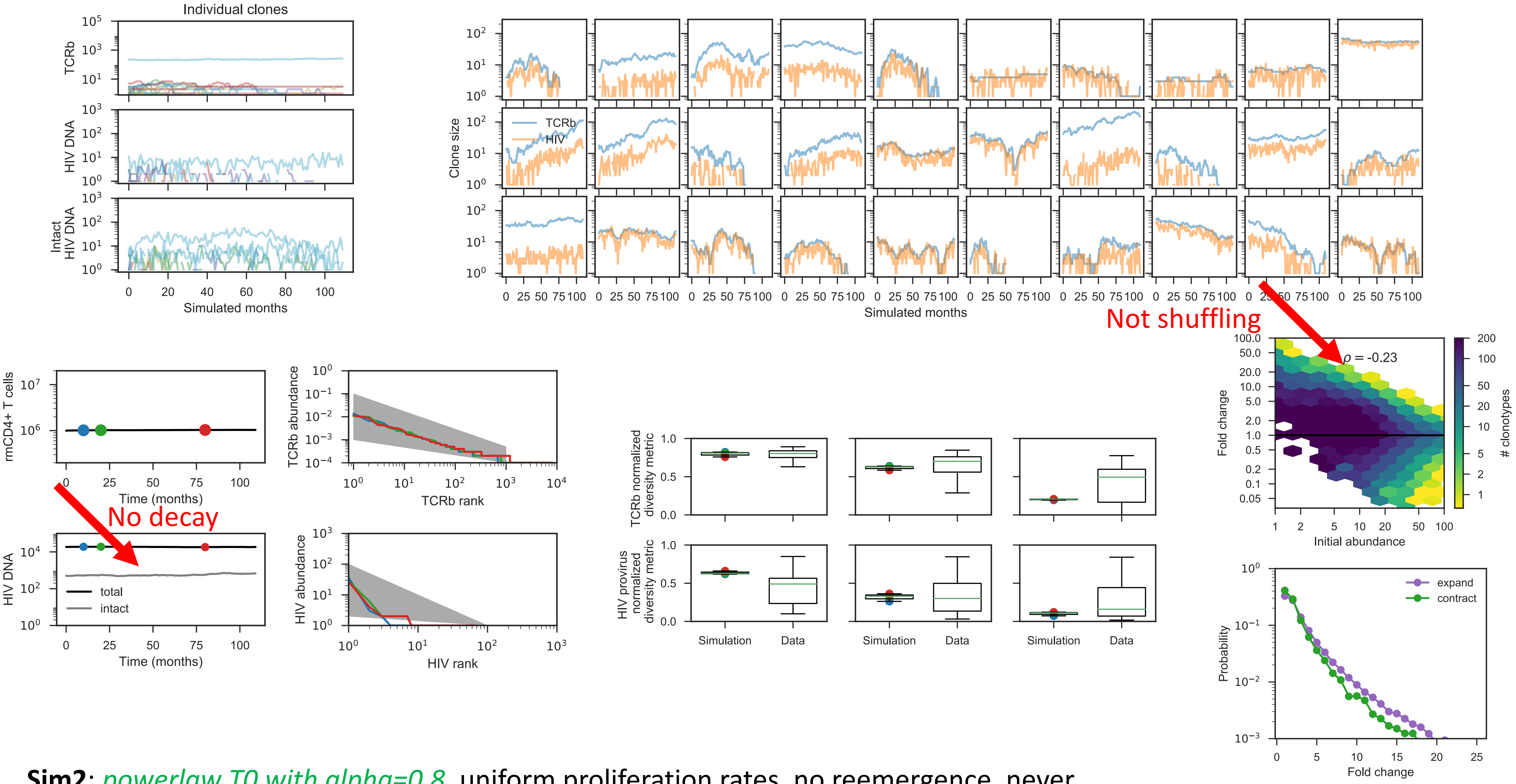


**Sim2**: *powerlaw T0 with alpha=0.8*, uniform proliferation rates, no reemergence, never redraw rates, no selection on proviruses, *random coverage of TCR by proviruses*

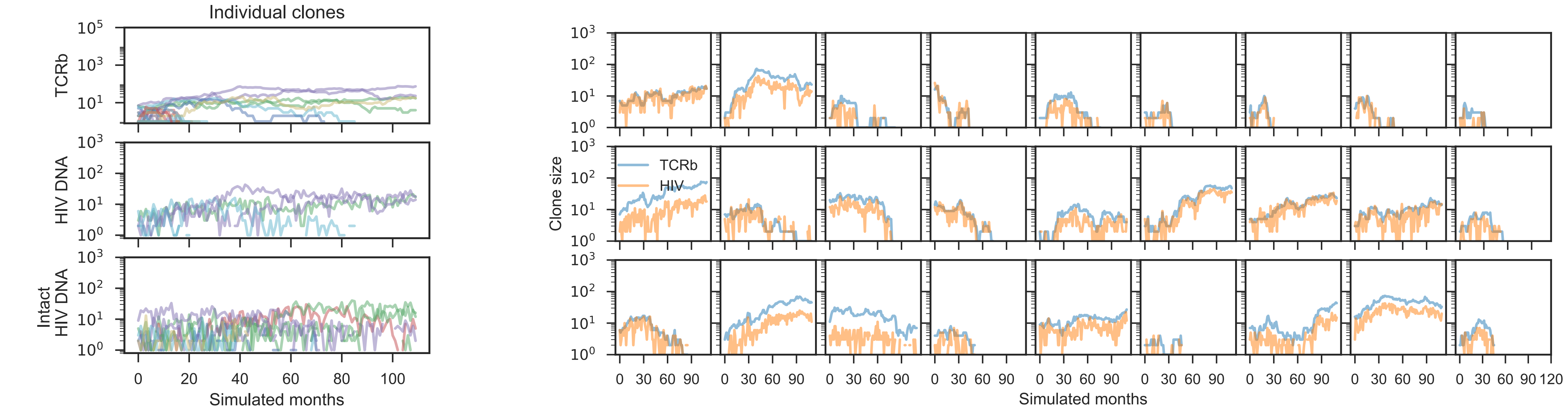


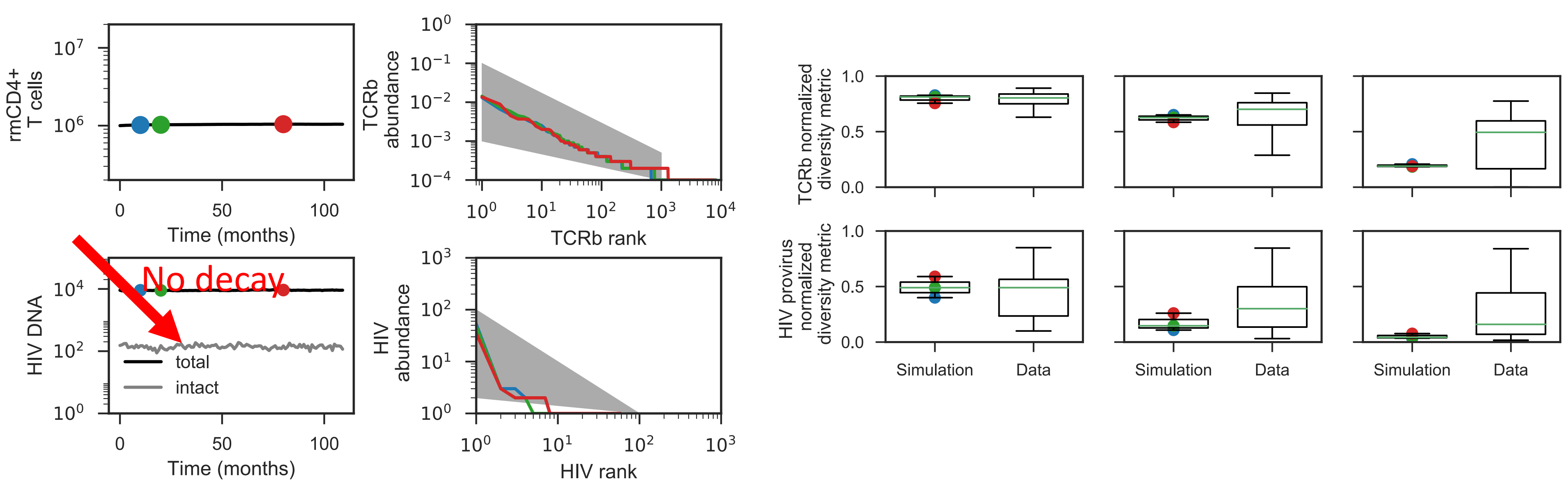


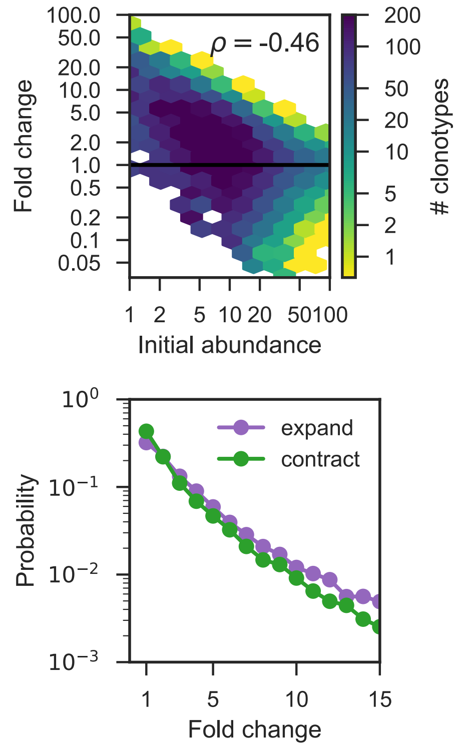


**Sim4**: *powerlaw T0 with alpha=0.8*, uniform proliferation rates, no reemergence, *redraw rates every 100 days,* no selection on proviruses, *random coverage of TCR by proviruses*

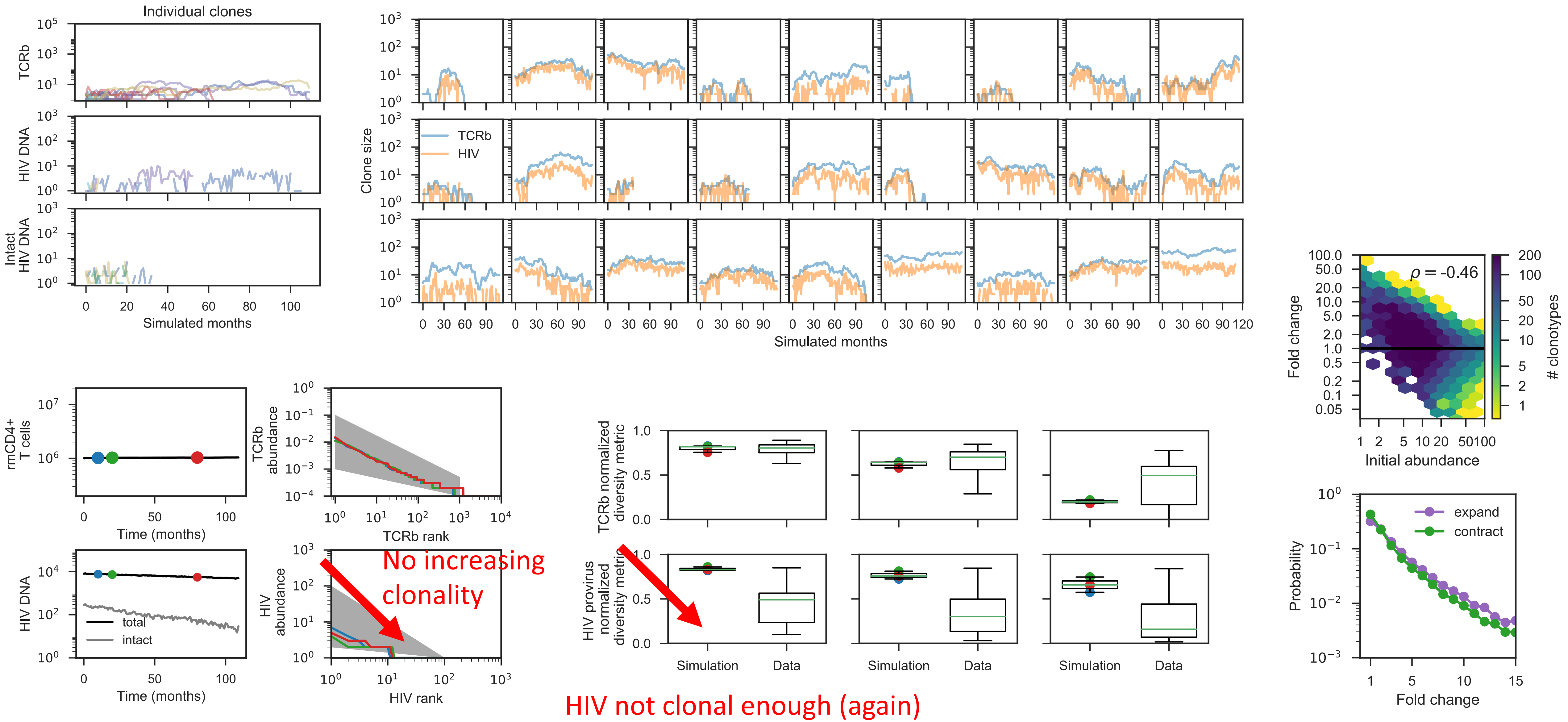


**Sim5**: *powerlaw T0 with alpha=0.8*, uniform proliferation rates, no reemergence, *redraw rates every 6mo*, *selection on intact proviruses (32mo half-life)*, *random coverage of TCR by proviruses*

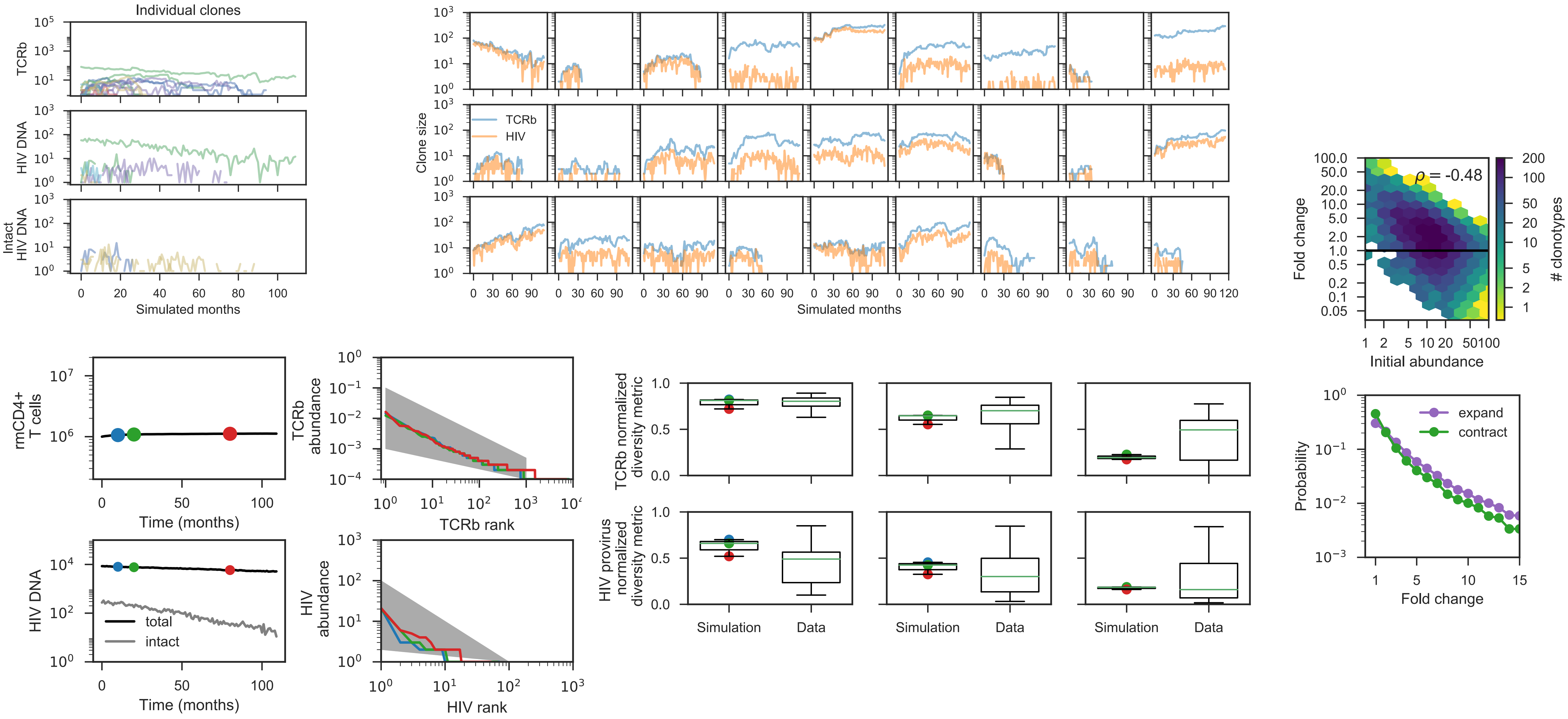


**Sim6**: *powerlaw T0 with alpha=0.8*, *exp proliferation rates (lam=1)*, no reemergence, *redraw rates every 6mo*, *selection on intact proviruses (32mo half-life)*, *random coverage of TCR by proviruses*
